# The urinary-metabolite-based lung cancer index (uLCI): an interpretable machine-learning risk model for early-stage disease

**DOI:** 10.64898/2026.06.26.26356700

**Authors:** Mohammed A Khan, Sharon R Pine, Frank J Gonzalez, Xin W Wang, Curtis C Harris, Daxesh P Patel

**Author notes:** Corresponding author Correspondence to: Curtis C. Harris, Laboratory of Human Carcinogenesis, Center for Cancer Research, National Cancer Institute, Bethesda, MD, USA and Daxesh P Patel, Laboratory of Human Carcinogenesis, Center for Cancer Research, National Cancer Institute, National Institutes of Health, Building 37, Room 3060, 37 Convent Drive, Bethesda, MD 20892, USA).

## Abstract

**Background:** Five-year survival from lung cancer exceeds 60% at stage I–II but falls below 10% once metastasis occurs. Low-dose CT (LDCT) screening reduces mortality in heavy smokers but carries a false-positive rate of approximately 29% and is restricted to smoking-based eligibility, leaving most cases undetected. We aimed to develop and independently validate an interpretable machine-learning urinary metabolite risk index (uLCI) for non-invasive lung cancer detection.

**Methods:** Four urinary metabolites—creatine riboside (CR), N-acetylneuraminic acid (NANA), 27-nor-5β-cholestane-3α,7α,12α,24*R*,25*S*-pentol (CP), and cortisol sulfate (CS)—and three clinical variables (age, race, smoking) were integrated by Lasso-regularised logistic regression into a uLCI score. The model was developed under 10-fold cross-validation in the NCI-Maryland (NCI-MD) cohort (n=845; 470 controls, 375 cases, stages I–IV) and applied without refitting to the independent Colorado Lung Cancer Cohort (n=488; 211 controls, 277 cases). Analyses were prespecified; reporting followed TRIPOD+AI.

**Findings:** uLCI achieved an area under the curve (AUC) of 0·906 (95% CI 0·887–0·926) in NCI-MD and 0·748 (0·701–0·793) in the independent Colorado cohort. Scores rose monotonically across stages in both cohorts (Spearman ρ=0·69 and 0·45; both p<0·0001). Stage-specific discrimination was preserved from stage I to IV (NCI-MD 0·900–0·927; Colorado 0·722–0·843). Net reclassification improvement over clinical variables was 1·24 (1·14–1·36) and 0·74 (0·56–0·90). uLCI tertiles stratified post-resection survival in stage I–II disease (adjusted hazard ratio 2·03, 1·26–3·27).

**Interpretation:** uLCI is an independently validated, interpretable urinary risk index that detects lung cancer across all stages, with monotonic stage progression and post-resection prognostic value. Its false-positive rate compares favourably with published estimates for LDCT and cell-free-DNA assays, supporting prospective head-to-head evaluation as a non-invasive triage tool, including in screening-ineligible populations.

**Research in context:** *Evidence before this study:* We searched PubMed, Embase, and Web of Science from Jan 1, 2000, to Jan 31, 2026, without language restriction, for biomarker-based diagnostic or risk models for lung cancer, using “lung cancer”, “early detection”, “biomarker”, “urine”, “metabolite”, “machine learning”, “TRIPOD”, and “validation in an independent cohort”. Five-year survival exceeds 60% at stage I–II but falls below 10% after metastasis. Low-dose CT reduces mortality in heavy smokers but carries an approximately 29% false-positive rate and excludes never-smokers, who account for 10–25% of US lung cancers and up to 40% globally; blood-based cell-free DNA and methylation assays report rates near 27%. Most prior urinary-metabolite work, including our 2024 creatine-riboside and N-acetylneuraminic-acid report, used case–control designs without a locked, integrated model, and—even where two cohorts were analyzed—lacked prespecified independent validation or TRIPOD+AI-compliant reporting; an interpretable urinary index integrating an expanded metabolite panel with clinical variables under these standards had not been described.

*Added value of this study:* Across two cohorts (1333 individuals), we developed and independently validated uLCI, an interpretable Lasso-regularised logistic index combining four urinary metabolites (CR, NANA, CP, CS) with age, race, and smoking, reported to TRIPOD+AI standards. The prespecified locked model achieved an AUC of 0·906 (95% CI 0·887–0·926) in NCI-MD development and 0·748 (0·701–0·793) in the independent Colorado cohort without refitting, with discrimination preserved from stage I to IV (0·900–0·927; 0·722–0·843). uLCI rose monotonically with stage in both cohorts (Spearman ρ=0·693 and 0·449; both p<0·0001), improved reclassification over clinical variables (net reclassification improvement 1·24 and 0·74), and independently stratified post-resection survival in stage I–II disease (adjusted hazard ratio 2·03 in NCI-MD; 3·81 in Colorado). On indirect benchmarking, the 17–19% false-positive rate was below published estimates for low-dose CT (∼29%) and DELFI (∼27%), and discrimination was preserved in never-smokers — whom low-dose CT excludes — and across racial subgroups in the diverse development cohort.

*Implications of all the available evidence:* A locked, independently validated, non-invasive urinary index that is interpretable, accurate from stage I, stage-responsive, and prognostic after resection addresses a defined detection gap, with a plausible role as a low-cost triage test that raises pre-test probability before imaging and extends risk assessment to screening-ineligible never-smokers, complementary to low-dose CT. Prospective screening-cohort evaluation (planned in the NCI PLCO and Southern Community Cohort biobanks), head-to-head comparison with blood-based assays, recalibration to screening prevalence, and replication in diverse cohorts are the warranted next steps.

## Introduction

Lung cancer remains the leading cause of cancer-related mortality worldwide, accounting for 1·80 million deaths in 2020.^1^ Five-year survival exceeds 60% for stage I–II disease but falls below 10% once distant metastasis is present, establishing stage at detection as the principal determinant of outcome.^2^ Annual low-dose CT (LDCT) screening in high-risk smokers reduces lung-cancer-specific mortality by 20–24% in randomized trials,^2–4^ yet two constraints limit its global reach: a false-positive rate of approximately 29%,^5,6^ and smoking-based eligibility criteria that exclude never-smokers, who account for 10–25% of lung cancers in the USA and up to 40% worldwide.^7–10^

Non-invasive biomarker assays could address both constraints. Established lung cancer risk-prediction tools, including PLCOm2012,^11^ rely on epidemiological inputs alone, while emerging blood-based assays—plasma cell-free DNA fragmentomics^12^ and multi-cancer early-detection panels^13^—report false-positive rates comparable to LDCT and stage-dependent detection that remains weakest in early-stage disease. A non-invasive biomarker that couples robust discrimination with interpretable machine-learning modelling, independently validated performance, and a decentralized analytical workflow would meaningfully advance early detection.

Urinary creatine riboside (CR) and N-acetylneuraminic acid (NANA) are tumor-associated metabolites with established diagnostic value in lung cancer across both never-smokers and ever-smokers.^14–16^ The present study extends this two-metabolite foundation in five ways: two additional metabolites (27-nor-5β-cholestane-3α,7α,12α,24*R*,25*S*-pentol, CP; cortisol sulfate, CS) are integrated into a four-metabolite panel; metabolites are combined with clinical variables in an interpretable machine-learning risk index (uLCI); reporting is fully TRIPOD+AI-compliant,^17^ including bootstrap 95% CIs, calibration analyses, decision-curve analysis, and net reclassification improvement; discrimination is reported stage-by-stage across all stages; and post-resection survival is stratified by Kaplan–Meier and Cox proportional-hazards analysis.

We hypothesized that uLCI would discriminate lung cancer from controls in both a development and an independent validation cohort, exhibit monotonic stage progression as a quantitative indicator of disease burden, retain prognostic value after surgical resection, perform equitably across racial and smoking subgroups, and satisfy TRIPOD+AI reporting standards. uLCI is positioned as a non-invasive triage tool to identify individuals at elevated probability of lung cancer for confirmatory imaging—complementary to, rather than competitive with, LDCT screening.

## Methods

### Study design, cohorts, and urinary metabolite measurement

This two-cohort biomarker development and independent validation study was approved by the Institutional Review Boards of the National Cancer Institute (protocol #05-C-N021) and the University of Maryland, Baltimore, and by the Colorado Multiple Institutional Review Board (#23-0941) and the University of Colorado Cancer Center PRMS Expedited Review Committee (PRMS 23-110), with written informed consent from all participants (Table 1), in accordance with the Declaration of Helsinki. Reporting follows the TRIPOD+AI checklist^17^ (appendix). The NCI-Maryland (NCI-MD) development cohort prospectively enrolled patients with histologically confirmed lung cancer (stages I–IV) and population-based controls from the greater Baltimore metropolitan region; the independent Colorado Lung Cancer Cohort is a prospectively-collected biobanked cohort at the University of Colorado. In the Colorado data file, four participants with ambiguous case-control labels were resolved using the lowercase status variable (three controls, one case), yielding the final analytic set of 488 (211 controls, 277 cases; appendix Table S1, Figure S1). Spot urine was collected at study entry before treatment, and four metabolites (CR, NANA, CP, CS) were quantified by targeted LC-MS/MS with stable-isotope internal standards; complete chromatography and MRM parameters are in appendix Table S2.^18^

**Table 1.** Stage-specific and subgroup discrimination of uLCI in development and independent validation cohorts.

| Cohort / subgroup | n (controls / cases) | AUC (95% CI) |
| --- | --- | --- |
| NCI-MD overall (10-fold CV) | 470 / 375 | 0.906 (0.887–0.926) |
| Stage I vs all controls | 470 / 182 | 0.900 (0.869–0.920) |
| Stage II vs all controls | 470 / 45 | 0.905 (0.862–0.938) |
| Stage III vs all controls | 470 / 79 | 0.927 (0.894–0.951) |
| Stage IV vs all controls | 470 / 58 | 0.925 (0.886–0.953) |
| Early (I+II) vs controls | 470 / 227 | 0.896 (0.872–0.918) |
| Late (III+IV) vs controls | 470 / 137 | 0.924 (0.900–0.948) |
| Colorado overall (independent) | 211 / 277 | 0.748 (0.701–0.793) |
| Stage I vs all controls | 211 / 98 | 0.722 (0.662–0.778) |
| Stage II vs all controls | 211 / 66 | 0.726 (0.656–0.792) |
| Stage III vs all controls | 211 / 35 | 0.843 (0.784–0.899) |
| Stage IV vs all controls | 211 / 69 | 0.784 (0.720–0.843) |
| Early (I+II) vs controls | 211 / 164 | 0.723 (0.669–0.773) |
| Late (III+IV) vs controls | 211 / 104 | 0.804 (0.754–0.854) |
| NCI-MD never-smokers | 203 / 114 | 0.94 (0.92–0.96) |
| NCI-MD ever-smokers | 267 / 261 | 0.88 (0.85–0.91) |
| Colorado never-smokers | 102 / 76 | 0.77 (0.69–0.84) |
| Colorado ever-smokers | 109 / 201 | 0.70 (0.63–0.76) |
| NCI-MD African American | 214 / 73 | 0.91 (0.87–0.94) |
| NCI-MD European American | 256 / 302 | 0.89 (0.86–0.92) |

### uLCI development

uLCI was developed by L1-regularised (Lasso) logistic regression integrating the four log-transformed, creatinine-normalized metabolite values with three clinical variables (age, race, smoking status), all standardized to NCI-MD-derived means and standard deviations. Regularization strength was selected over 20 candidate values by 10-fold cross-validated area under the curve (AUC). The decision threshold was the Youden-J optimum on NCI-MD cross-validated predictions, applied identically to Colorado without re-selection. The locked NCI-MD model was applied to Colorado with no re-fitting, re-tuning, or threshold adjustment; the Colorado cohort was used solely for independent validation. Full model coefficients and the prediction formula are in appendix Table S3; the analytical pipeline is detailed in appendix Section S1.

### Statistical analysis

Discrimination was summarized by AUC with 95% CIs from 2000-iteration bootstrap. Incremental value over clinical variables was quantified by category-free net reclassification improvement (NRI) and integrated discrimination improvement (IDI).^19^ Stage progression was tested by Spearman correlation across ordinal stage (0=control, 1–4=cases). Calibration was assessed by Hosmer–Lemeshow test, calibration plot, and Brier score, with intercept–slope recalibration (Steyerberg method).^20^ Net clinical benefit was assessed by decision-curve analysis.^21^ Survival was analyzed by Kaplan–Meier estimation with uLCI tertiles and three-group log-rank test; Cox proportional-hazards regression assessed prognostic contribution adjusted for age, sex, race, and smoking. Analyses used R 4·2·0, with glmnet for Lasso-regularised logistic regression, pROC for discrimination, boot for bootstrap confidence intervals, ResourceSelection for Hosmer–Lemeshow calibration, dcurves for decision-curve analysis, nricens for net reclassification and integrated discrimination improvement, caret for benchmark classifiers and permutation importance, and survival with survminer for time-to-event analyses; a two-sided p<0·05 was considered significant.

## Results

### uLCI rises monotonically with stage

In NCI-MD (845 participants; 470 controls, 375 cases across stages I–IV), median uLCI was 0·15 in controls, 0·75 in stage I (n=182), 0·70 in stage II (n=45), 0·86 in stage III (n=79), and 0·91 in stage IV (n=58), with strong monotonic stage progression (Spearman ρ=0·69, p<0·0001; Figure 1A). The pattern replicated in the independent Colorado validation cohort (488 participants; 211 controls, 277 cases): median uLCI 0·34 in controls, 0·68 (stage I, n=98), 0·71 (stage II, n=66), 0·81 (stage III, n=35), and 0·78 (stage IV, n=69; ρ=0·45, p<0·0001; Figure 1B). The full per-stage probability-density distributions, restricted to cases, are shown in appendix Figure S2. The monotonic relationship across two independent cohorts—each spanning the full clinical stage spectrum—indicates that uLCI quantifies lung-cancer-associated metabolic perturbation rather than a binary classification signal.

**Figure 1:**
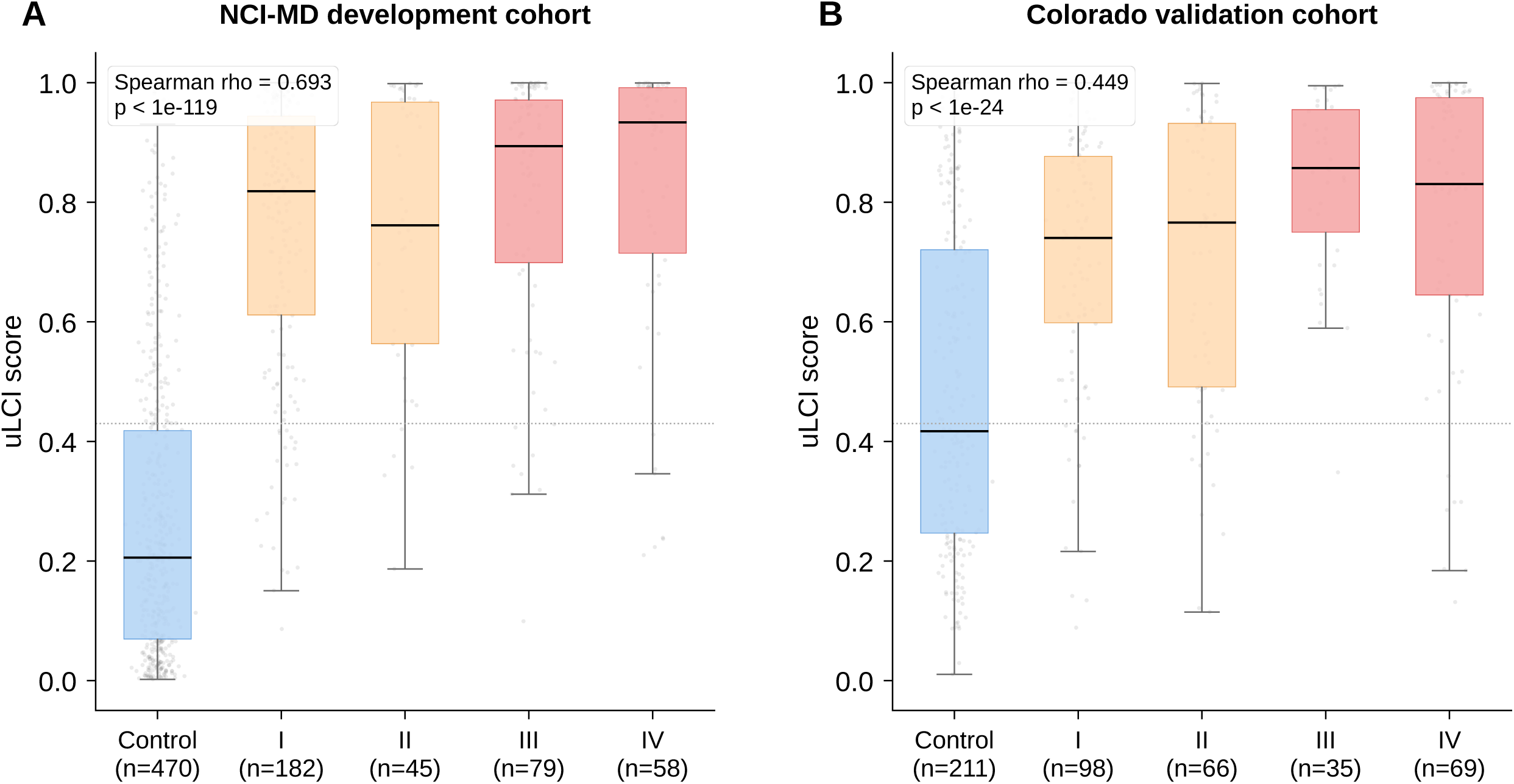
uLCI score distributions by case-control status and stage. (A) NCI-MD development cohort. (B) Colorado independent validation cohort. Boxplots show median (line), interquartile range (box), and 1·5×IQR whiskers; outliers shown as points. uLCI rises monotonically with stage in both cohorts (Spearman ρ=0·693 NCI-MD, p<0·0001; ρ=0·449 Colorado, p<0·0001). Dotted line indicates the Youden-J-optimised decision threshold (0·43) selected on NCI-MD cross-validated predictions and applied identically to Colorado without re-selection.

### uLCI discriminates lung cancer across all stages

Stage-specific discrimination compared each disease stage against the full control set within each cohort (Figure 2). In NCI-MD, AUCs were preserved across stages: stage I 0·900 (95% CI 0·869–0·920), stage II 0·905 (0·862–0·938), stage III 0·927 (0·894–0·951), and stage IV 0·925 (0·886–0·953; Figure 2A). In Colorado, the pattern replicated under locked-model independent validation: stage I 0·722 (0·662–0·778), stage II 0·726 (0·656–0·792), stage III 0·843 (0·784– 0·899), and stage IV 0·784 (0·720–0·843; Figure 2B). The development-cohort AUC across all stages was 0·906 (0·887–0·926); the locked NCI-MD model applied to Colorado achieved 0·748 (0·701–0·793). A clinical-variables-only model achieved 0·632 (0·595–0·668) in NCI-MD and 0·625 (0·581–0·669) in Colorado (ΔAUC 0·27 development; 0·12 validation; bootstrap p<0·0001 for both). NRI was 1·24 (1·14–1·36) in NCI-MD and 0·74 (0·56–0·90) in Colorado; IDI was 0·44 (0·41–0·48) and 0·23 (0·18–0·27), respectively (appendix Figure S3).

**Figure 2:**
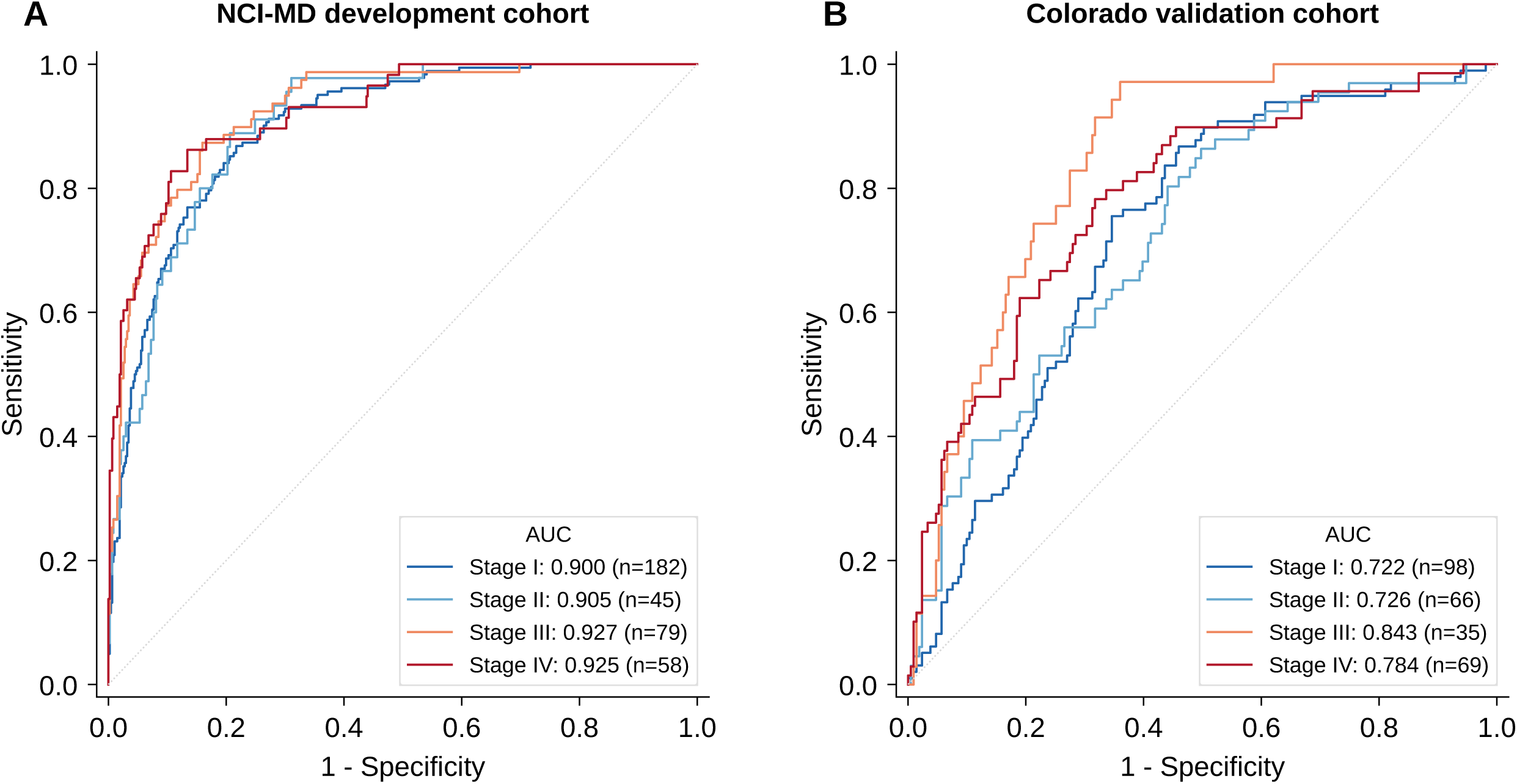
Stage-stratified receiver-operating-characteristic curves: uLCI discrimination of each disease stage versus all controls. (A) NCI-MD development cohort. (B) Colorado independent validation cohort. AUCs with bootstrap 95% CIs (2000 resamples) are shown for each stage. uLCI maintains preserved discrimination at stage I (NCI-MD AUC 0·900; Colorado 0·722)—the stage at which intervention has the largest impact on mortality.

### Performance across racial and smoking subgroups

Subgroup analyses prespecified by race and smoking status are in Table 1. Discrimination was preserved across racial subgroups in the diverse NCI-MD cohort (African American AUC 0·91, 95% CI 0·87–0·94, n=287; European American 0·89, 0·86–0·92, n=558). The Colorado African American subgroup (n=18) was too small for meaningful stratified analysis—a priority for replication. Performance was preserved across smoking strata (never-smokers: NCI-MD 0·94, Colorado 0·77; ever-smokers: NCI-MD 0·88, Colorado 0·70), with never-smoker performance equal to or stronger than ever-smoker performance, supporting potential applicability in populations currently outside LDCT-screening eligibility (appendix Figure S4).

### Calibration and prognostic stratification of survival

uLCI was well-calibrated in NCI-MD (Hosmer–Lemeshow p=0·052; Brier score 0·124). In Colorado, initial calibration on the absolute-probability scale was poor (Hosmer–Lemeshow p<0·001); intercept–slope recalibration (slope 0·524, intercept correction +0·075) improved Hosmer–Lemeshow to p=0·033 without altering AUC or rank-ordering (appendix Figure S5). Decision-curve analysis demonstrated positive net clinical benefit across plausible threshold probabilities in both cohorts. In stage I–II disease with available post-resection follow-up, uLCI tertiles stratified overall survival in both cohorts (Figure 3): NCI-MD three-group log-rank p=0·036 (n=227); Colorado p=0·001 (n=164). Adjusted for age, sex, race, and smoking, the high-vs-low tertile hazard ratio was 2·03 (95% CI 1·26–3·27, p=0·004) in NCI-MD and 3·81 (1·61– 9·02, p=0·002) in Colorado (appendix Figure S6, Table S4).

**Figure 3:**
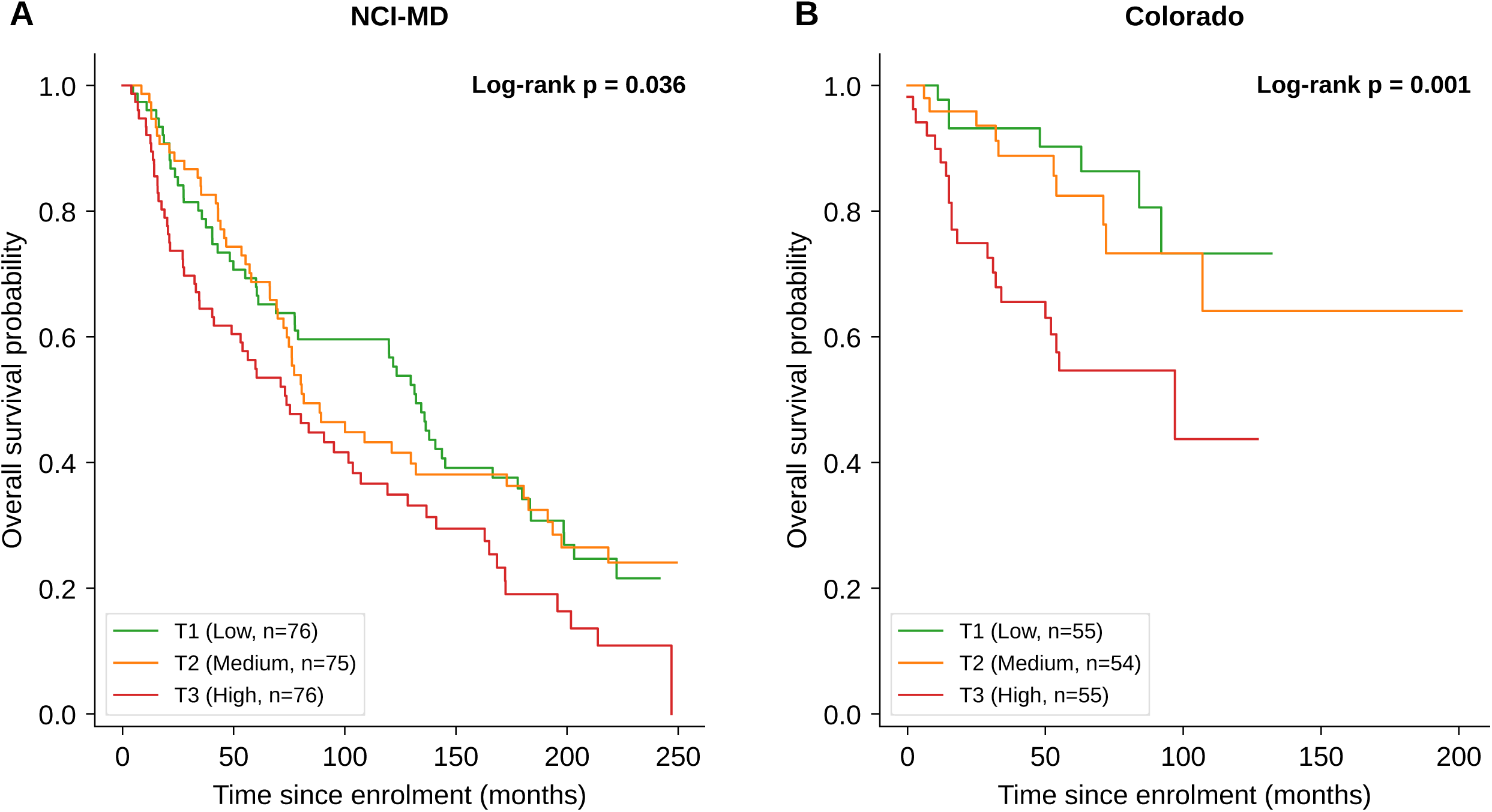
Kaplan–Meier overall-survival curves stratified by within-cohort uLCI tertile in stage I–II cases with post-resection follow-up. (A) NCI-MD development cohort (n=227): 3-group log-rank p=0·036; Cox-adjusted high-vs-low tertile HR 2·03 (95% CI 1·26–3·27, p=0·004). (B) Colorado validation cohort (n=164): 3-group log-rank p=0·001; Cox-adjusted high-vs-low tertile HR 3·81 (1·61–9·02, p=0·002). Cox models adjusted for age, sex, race, and smoking status. Cause-specific death information was not available in either cohort, so competing-risk (Fine-Gray) analysis was not feasible; all-cause-mortality Cox analysis is reported.

## Discussion

In two independent cohorts totalling 1333 individuals, we developed and independently validated uLCI, an interpretable machine-learning urinary metabolite risk index for lung cancer. The model was developed and locked entirely within the NCI-MD cohort and then applied, without any refitting, retuning, or threshold reselection, to the independent Colorado cohort, whose urine samples were assayed by the identical LC-MS/MS protocol. uLCI achieved an AUC of 0·906 (95% CI 0·887–0·926) in NCI-MD and 0·748 (0·701–0·793) on locked-model independent validation in Colorado. The principal finding is the monotonic increase of uLCI with stage in both cohorts (Spearman ρ=0·69 in NCI-MD and 0·45 in Colorado; both p<0·0001), with preserved stage-specific discrimination from stage I (AUC 0·90 development; 0·72 validation) to stage IV (0·92; 0·78). Net reclassification improvement over clinical variables alone was large in both cohorts (1·24 in NCI-MD; 0·74 in Colorado), and uLCI tertiles stratified post-resection overall survival in stage I–II disease (adjusted high-vs-low hazard ratio 2·03 in NCI-MD; 3·81 in Colorado).

The principal clinical implication is that a non-invasive urinary score can identify individuals with elevated probability of lung cancer across all clinically actionable stages, with discrimination preserved at stage I—the stage where intervention has the largest effect on mortality. uLCI is not proposed as a replacement for LDCT but as a complementary, low-cost, decentralized first-line evaluation that may improve downstream screening efficiency by raising the pre-test probability of disease. The monotonic stage progression is notable: uLCI behaves as a quantitative indicator of disease burden, suggesting utility beyond binary triage, as a stratification variable in screening, surveillance, or adjuvant-therapy decisions.^22^

uLCI’s stage I performance is its most clinically consequential property, because this is precisely where established blood-based assays are weakest. Circulating tumour DNA mutation-calling approaches report sensitivities as low as 40% for early-stage lung cancer, constrained by the extremely low tumor-derived DNA fraction shed by stage I tumors.^12^ Methylation- and fragmentomic-based cfDNA approaches partially mitigate this but report stage I sensitivities of approximately 25–60%, generally insufficient for stand-alone early detection. uLCI is mechanistically distinct: rather than depending on the quantity of tumour-shed nucleic acid in plasma, it measures stable, abundant small-molecule metabolites concentrated in urine, decoupling detection from circulating tumour fraction. This may explain the preserved stage I discrimination observed here and positions a urinary LC-MS/MS metabolite panel as an orthogonal, complementary modality to plasma cfDNA assays—potentially additive within a combined screening pathway. Beyond nucleic-acid assays, blood-based metabolite panels measured by LC-MS/MS have also been proposed for early lung cancer detection, but report stage-dependent performance and have not been locked and validated in an independent cohort under TRIPOD+AI reporting;^23,24^ uLCI differs in using a non-invasive urinary matrix, an interpretable locked model, and stage-resolved, independently validated discrimination. The 17– 19% false-positive rate compares favorably with published estimates for LDCT (approx. 29%), plasma cfDNA fragmentomics (approx. 27%),^12^ and methylation-based multi-cancer early-detection panels, although head-to-head evaluation on the same participants remains the definitive test.

The development-to-validation AUC difference (0·906 to 0·748) warrants explicit interpretation. All four metabolite baseline concentrations were higher in Colorado controls than in NCI-MD controls (all Wilcoxon p<0·0001), shifting the decision boundary and compressing case–control separation; case-versus-control effect sizes (Cohen’s d on log-transformed values) were attenuated by 36% for CR, 64% for NANA, 51% for CP, and 20% for CS (appendix Figures S7– S8). Three population-level factors contribute: first, an 8·5-fold reduction in African American representation (34% to 4%) means the race coefficient (β=–0·55) learned in NCI-MD adds noise rather than signal in Colorado; second, Colorado controls were 3·1 years older with a 20% wider age SD (11·0 vs 9·2), increasing overlap between age-related metabolic variation and the cancer signal; and third, the higher case prevalence in Colorado (57% vs 44%) shifts the calibration intercept and accounts for the recalibration slope of 0·524. These sources of population shift are characteristic of stringent locked-model independent validation and do not indicate model failure: the preserved rank-ordering (AUC 0·748 remains significantly above the clinical-variables-only baseline of 0·625; bootstrap p<0·0001) and the replicated monotonic stage progression (ρ=0·45) confirm that the biological signal is transportable across populations. The attenuated stage correlation in Colorado (ρ=0·45 vs 0·69 in NCI-MD) reflects the same population shift rather than a distinct phenomenon: compression of case–control separation reduces the score’s dynamic range across stages, and Colorado’s stage-frequency distribution differs materially (stage I 35% vs 49%; stage IV 25% vs 15%), with a non-monotonic median uLCI at stage IV (0·78, below stage III’s 0·81) that mechanically lowers a rank-based coefficient even as the overall positive trend remains highly significant (p<0·0001). Site-specific recalibration, as demonstrated here, is a routine pre-deployment step for any clinical prediction model applied to a new population.

Four methodological strengths merit emphasis. First, the locked, non-refitted independent validation in a separate cohort under population shift is a stringent test of generalizability that many biomarker studies omit. Second, bootstrap 95% CIs on all discrimination metrics, calibration with formal recalibration, decision-curve analysis, and net reclassification improvement together meet the TRIPOD+AI reporting standard.^17^ Third, uLCI uses interpretable machine learning—a sparse, regularized generalized linear model^25^ whose coefficients are transparent and reportable; on a fixed seven-variable feature space, deep learning is not expected to outperform regularized regression on tabular data of modest dimensionality **(appendix Table S5)**.^26,27^ Fourth, the diverse racial composition of NCI-MD allowed direct evaluation of cross-racial performance—a health-equity consideration often omitted from biomarker studies^28^ —and performance was preserved across racial subgroups and across never-smokers and ever-smokers, suggesting the urinary signal does not depend on smoking-induced metabolic perturbation.^29^

Six limitations should be acknowledged. First, both cohorts are case-control rather than prospective screening cohorts; confirmation in true screening populations is essential before deployment and is being pursued through planned analyses in the NCI PLCO and Southern Community Cohort biobanks. Second, head-to-head comparison with established blood-based assays on the same samples was not possible; comparative claims rest on published performance estimates. Third, Colorado calibration required intercept–slope recalibration (slope 0·524, intercept correction +0·075), consistent with the documented population shift and reflecting the expected behavior of a locked model applied without refitting to a demographically distinct population rather than intrinsic instability. Fourth, the racial mismatch between NCI-MD (34% African American) and Colorado (4%) contributes to both the AUC gap and the calibration shift; replication of African American subgroup performance in larger diverse independent cohorts is a priority. Fifth, Colorado smoking status was recorded as free text and harmonized to a binary never/ever variable using prespecified keyword rules with senior-author audit; the full mapping is provided in the appendix (Table S6). Sixth, the biological mechanism underlying CP’s association with lung cancer is not yet established and is under separate investigation.

In conclusion, uLCI is an independently validated, interpretable machine-learning urinary risk index for non-invasive lung cancer detection, with monotonic stage progression and preserved performance across racial subgroups and smoking strata. The assay is non-invasive, analyte-stable, and CLIA-compatible, and is positioned as a complementary triage tool to LDCT screening and to plasma cfDNA assays. Prospective evaluation in screening populations, head-to-head comparison with blood-based assays on the same samples, recalibration for screening-prevalence settings, and pan-cancer extension are the warranted next steps.

## Supporting information

Completed TRIPOD+AI reporting checklist

## Data Availability

All data produced in the present work are contained in the manuscript and its supplementary materials. The raw data were generated at the National Cancer Institute, National Institutes of Health, and are available from the corresponding author upon reasonable request.

## Contributors

MAK conceptualized the study and performed the statistical and computational analyses. SRP provided Colorado cohort samples. CCH provided study materials and resources. MAK and DPP drafted the manuscript. SRP, FJG, CCH, and XWW critically reviewed, edited, and approved the manuscript. CCH supervised the study. DPP developed and performed the original analytical assays and is the corresponding author. MAK and DPP directly accessed and verified the reported underlying data. All authors provided their final approval of the version to be published. All authors accepted responsibility to submit the manuscript for publication. All authors had full access to the study data and took responsibility for its integrity and accuracy.

## Data sharing

De-identified individual participant data underlying the reported analyses are available subject to NCI IRB approval and a data-use agreement on reasonable request to the corresponding author. Analytical code will be deposited on Zenodo upon acceptance with a DOI.

## Acknowledgments

We thank participants and their families, and the research and clinical staff of the NCI-MD lung cancer case-control study and the Colorado Lung Cancer Cohort. This work was supported by the Intramural Research Program of the U.S. National Institutes of Health (NIH), National Cancer Institute, Center for Cancer Research (grant ZIA BC 011492). The funding body played no role in the design and conduct of the study; collection, management, analysis, and interpretation of the data; preparation, review, or approval of the manuscript; or the decision to submit the manuscript for publication. The contributions of the NIH author(s) were made as part of their official duties as NIH federal employees, are in compliance with agency policy requirements, and are considered Works of the United States Government. However, the findings and conclusions presented in this paper are those of the authors and do not necessarily reflect the views of the NIH or the U.S. Department of Health and Human Services.

## Use of generative AI and AI-assisted technologies

During the preparation of this manuscript, the authors used Claude Sonnet 4.6 (Anthropic) for grammar and language editing only. After using this tool, the authors reviewed and edited the content as needed and take full responsibility for the content of the publication.

