## Supplemental Data 1 for "The urinary-metabolite-based lung cancer index (uLCI): an interpretable machine-learning risk model for early-stage disease"

Mohammed A Khan, [Sharon R Pine](https://pubmed.ncbi.nlm.nih.gov/?term=%22Pine%20SR%22%5bAuthor%5d), [Frank J Gonzalez](https://pubmed.ncbi.nlm.nih.gov/?term=%22Gonzalez%20FJ%22%5bAuthor%5d), Curtis C Harris, Xin W Wang, Daxesh P Patel

**Contents**

Section S1. Detailed methods (full LC-MS/MS, Lasso derivation, statistical pipeline)

Table S1. Baseline demographic and clinical characteristics of both cohorts

Table S2. Urine sample preparation, targeted LC-MS/MS chromatography, and MRM parameters

Table S3. Locked uLCI Lasso coefficients on standardized features

Table S4. Cox proportional-hazards analysis for individual urinary metabolites

Table S5. Benchmark classifier comparison under identical 10-fold cross-validation

Table S6. Colorado smoking status mapping rules (14 free-text entries → binary)

Figure S1. STARD-style participant flow diagrams for both cohorts

Figure S2. uLCI predicted probability distributions by stage (both cohorts)

Figure S3. Overall ROC curves: uLCI vs clinical-only model (both cohorts)

Figure S4. uLCI score distributions by smoking status and case-control group

Figure S5. Colorado calibration before and after intercept-slope recalibration

Figure S6. Overall survival by uLCI tertile — all stages combined (both cohorts)

Figure S7. Individual urinary metabolite distributions in NCI-MD by smoking status

Figure S8. Individual urinary metabolite distributions in Colorado by smoking status

**Section S1. Detailed methods**

**S1·1 Urinary metabolite quantification**

Spot urine samples were collected at study entry, prior to surgical resection or systemic therapy initiation, aliquoted within 4 hours, and stored at –80 °C until analysis. Four metabolites (creatine riboside [CR], N-acetylneuraminic acid [NANA], cortisol sulfate [CS], and 27-nor-5β-cholestane-3α,7α,12α,24,25-pentol [CP]) were quantified by targeted LC-MS/MS with stable-isotope-labelled internal standards. CR and NANA were analyzed under hydrophilic interaction liquid chromatography (HILIC); CS and CP under reverse-phase chromatography. Complete chromatography and MRM parameters are in Table S2. All measurements were normalized to urinary creatinine. Stability over 7 days at room temperature and 4 °C was confirmed in a pilot subset, supporting decentralized collection workflows.

**S1·2 uLCI model derivation**

uLCI was developed as a sparse, interpretable, regularized generalized linear model. Each metabolite value was log-transformed (log1p), creatinine-normalized, and standardized to zero mean and unit variance using NCI-MD-derived parameters; the same standardization parameters were applied identically to Colorado without re-fitting. Three clinical variables (age continuous, race binary AA=1 vs EA=0, smoking binary ever=1 vs never=0) were similarly standardized.

uLCI is computed as:

uLCI = σ(β₀ + β₁·CR + β₂·NANA + β₃·CP + β₄·CS + β₅·age + β₆·race + β₇·smoking)

where σ is the logistic sigmoid function and all coefficients (β₀…β₇) are reported in Table S3.

The L1 regularization strength was selected over 20 candidate values by 10-fold cross-validated AUC. The decision threshold for binary classification was the Youden-J optimum on NCI-MD cross-validated predictions (threshold = 0·43) and applied identically to Colorado without re-selection. The locked NCI-MD model was applied to Colorado with no re-fitting, re-tuning, or threshold adjustment, in compliance with TRIPOD+AI guidance for external validation.

**S1·3 Statistical analysis**

Discrimination was summarized by AUC with 95% CIs from 2000-iteration bootstrap. Pairwise AUC comparisons (full vs clinical-only) used the same bootstrap framework with two-sided p-values derived from the proportion of paired AUC differences crossing zero. Incremental value over clinical variables was further quantified by category-free Net Reclassification Improvement (NRI) and Integrated Discrimination Improvement (IDI). Classification metrics (accuracy, sensitivity, specificity, PPV, NPV, Cohen's κ, false-positive rate) were reported at the prespecified Youden-J threshold. Stage progression was tested by Spearman rank correlation across ordinal stage (0=control, 1-4=cases). Calibration was assessed by Hosmer–Lemeshow goodness-of-fit test (10 deciles), calibration curve (10 quantile bins), and Brier score; intercept-slope recalibration followed Steyerberg method. Net clinical benefit was assessed by decision-curve analysis across plausible threshold probabilities. Survival was analyzed by Kaplan-Meier estimation with uLCI categorised by within-cohort tertiles, comparing strata by 3-group log-rank test; Cox proportional-hazards regression assessed prognostic contribution adjusted for age, sex, race, and smoking. Benchmark comparison with k-nearest neighbors, support vector machine, random forest, and naïve Bayes used identical 10-fold cross-validation on the same feature set (Table S5). Analyses were conducted in R 4·2·0 (glmnet, pROC, boot, ResourceSelection, dcurves, nricens, caret, survival, and survminer).

**Table S1. Baseline demographic and clinical characteristics**

| Characteristic | NCI-MD (n=845) | Colorado (n=488) |
| --- | --- | --- |
| Controls / cases, n | 470 / 375 | 211 / 277 |
| Cases by stage, n |  |  |
| Stage I | 182 (49%) | 98 (35%) |
| Stage II | 45 (12%) | 66 (24%) |
| Stage III | 79 (21%) | 35 (13%) |
| Stage IV | 58 (15%) | 69 (25%) |
| Stage missing | 11 (3%) | 9 (3%) |
| Smoking status, n |  |  |
| Never-smokers | 317 (38%) | 178 (36%) |
| Ever-smokers | 528 (62%) | 310 (64%) |
| Age, mean (SD), years | 66·1 (9·2) | 69·2 (11·0) |
| Female sex, n (%) | 425 (50%) | 217 (44%) |
| Race, n (%) |  |  |
| African American | 287 (34%) | 18 (4%) |
| European American | 558 (66%) | 446 (91%) |
| Other / Unknown | 0 (0%) | 24 (5%) |

NCI-MD is the development cohort; Colorado is the independent external validation cohort. Stage assigned using AJCC TNM criteria. Stage I-II represents early-stage disease; stage III-IV represents late-stage disease. The substantial racial composition difference between cohorts (NCI-MD 34% AA vs Colorado 4%) is identified as a limitation in the main text Discussion.

**Table S2. Targeted LC-MS/MS chromatography and MRM parameters**

Urine sample preparation and chromatography conditions for quantification of the four metabolites. Samples were processed by two complementary methods: HILIC phase for hydrophilic metabolites (CR, NANA) and reverse-phase (RP) for less-polar metabolites (CS, CP).

Sample preparation

HILIC phase: 60 µL urine + 300 µL extraction buffer with stable-isotope-labelled internal standards (1:6 dilution); centrifuged at 15,000 g for 15 min (recover 360 µL); 80 µL extract + 80 µL 75% acetonitrile without standards (160 µL total, 1:12 final dilution).

Reverse phase: 60 µL urine + 60 µL extraction buffer with internal standards (1:1 dilution); centrifuged at 15,000 g for 15 min (recover 360 µL); 100 µL transferred for injection (1:2 final dilution).

**Chromatography, MRM transitions, and internal standards**

| Chromatography phase, column, run time | Analyte | MRM transition (m/z); RT | Internal standard (MRM; RT; concentration) |
| --- | --- | --- | --- |
| HILIC UPLC BEH Amide 1·7 µm, 50 × 2·1 mm; 12 min | CR | 264·12 → 132·07; RT 3·4 min | CR-¹³C,¹⁵N₂ (267·12 → 134·97; RT 3·4 min; 0·2 µM) |
| HILIC UPLC BEH Amide 1·7 µm, 50 × 2·1 mm; 12 min | NANA | 310·74 → 274·06; RT 3·4 min | N-acetyl-D-[1,2,3-¹³C₃] neuraminic acid (313·12 → 277·26; RT 3·4 min; 20 µM) |
| RP UPLC BEH C18 1·7 µm, 50 × 2·1 mm; 12 min | CS | 443·07 → 363·16; RT 5·3 min | Hydrocortisone-9,11,12,12-d₄ 21-sulfate (447·07 → 367·16; RT 5·3 min; 10 µM) |
| RP UPLC BEH C18 1·7 µm, 50 × 2·1 mm; 12 min | CP | 403·33 → 385·33; RT 6·0 min | NCP-D4 (407·33 → 389·33; RT 6·0 min; 4 µM) |

CR=creatine riboside; NANA=N-acetylneuraminic acid; CS=cortisol sulfate; CP=27-nor-5β-cholestane-3α,7α,12α,24,25-pentol; MRM=multiple-reaction monitoring; RT=retention time; UPLC BEH=ultra-performance liquid chromatography with ethylene-bridged hybrid particles. All quantification was performed with stable-isotope-labelled internal standards. All metabolite values are normalized to urinary creatinine.

**Table S3. Locked uLCI Lasso coefficients**

Locked uLCI model coefficients on standardised features. The prediction is computed as σ(β·X + β₀), where σ is the logistic sigmoid, X is the seven-element standardised feature vector, and β₀ is the intercept. Features are standardised using NCI-MD parameters and applied identically to Colorado without re-fitting.

| Feature | Standardised β | Direction | Interpretation |
| --- | --- | --- | --- |
| CR | +1·27 | Positive | Higher CR → higher uLCI |
| NANA | +1·25 | Positive | Higher NANA → higher uLCI |
| CP | +0·98 | Positive | Higher CP → higher uLCI |
| CS | ≈0 | Near-zero | Lasso shrinkage near zero for diagnosis; however, CS shows independent prognostic signal (Colorado Cox HR 1·27, p=0·030; Table S4) and independent univariate diagnostic signal in the ever-smoker stratum, supporting retention |
| Age (continuous, standardised) | –0·24 | Negative | Age effect in NCI-MD; cohort-specific |
| Race (AA=1 vs EA=0) | –0·55 | Negative | Reflects NCI-MD composition |
| Smoking status (ever=1 vs never=0) | +0·39 | Positive | Smoking adds modest risk increment |
| Intercept (β₀) | +0·15 | — | Calibrated to NCI-MD prevalence |

CR, NANA, and CP carry the principal discriminative weight. CS contributes near-zero weight after Lasso regularization but is retained in the panel as it provided independent univariate signal in the ever-smoker stratum. Negative race and age coefficients reflect within-cohort case-control composition, not true biological direction.

**Table S4. Cox proportional-hazards analysis for individual urinary metabolites**

Cox proportional-hazards regression for overall survival in cases, with each individual urinary metabolite entered as a standardized continuous predictor (per standard-deviation increase of log-transformed, creatinine-normalized value), adjusted for age, race, and smoking status. Hazard ratios above 1·00 indicate higher metabolite levels associated with worse survival. All-cause mortality is reported; cause-of-death information was not available in either cohort, so competing-risk (Fine–Gray sub-distribution hazard) analyses could not be performed and are deferred to future studies with cause-specific death data.

| Metabolite | NCI-MD cohort HR (95% CI), p | Colorado cohort HR (95% CI), p |
| --- | --- | --- |
| CR | 1·35 (1·25–1·45), p<0·0001 | 1·76 (1·49–2·07), p<0·0001 |
| NANA | 1·18 (1·09–1·28), p<0·0001 | 1·27 (1·03–1·56), p=0·025 |
| CP | 1·10 (1·01–1·19), p=0·032 | 1·02 (0·84–1·24), p=0·83 |
| CS | 1·07 (0·95–1·21), p=0·24 | 1·27 (1·02–1·58), p=0·030 |

NCI-MD: n=375 cases with survival data, 297 deaths; Colorado: n=277 cases with survival data, 91 deaths. All HRs are per 1-SD increase in the log-transformed, creatinine-normalised metabolite value, adjusted for age (continuous), race (AA vs EA), and smoking status (ever vs never). CR shows the strongest and most consistent prognostic signal across both cohorts; NANA shows significant association in both; CP and CS show cohort-specific significance. uLCI combines all four metabolites with the three clinical variables into a single integrated risk score.

**Table S5. Benchmark classifier comparison**

Benchmark classifier comparison under identical 10-fold cross-validation in the NCI-MD development cohort. All classifiers used the same seven standardized features (four metabolites + three clinical variables). Lasso logistic regression (used in the locked uLCI model) is shown in bold.

| Classifier | AUC (10-fold CV) | Sensitivity | Specificity | Interpretability |
| --- | --- | --- | --- | --- |
| Lasso logistic regression (uLCI) | 0·906 | 0·83 | 0·83 | High (sparse linear coefficients) |
| k-Nearest Neighbours (k=15) | 0·89 | 0·82 | 0·81 | Low (distance-based) |
| Support Vector Machine (RBF) | 0·92 | 0·84 | 0·84 | Low (non-linear kernel) |
| Random Forest (n=200) | 0·93 | 0·85 | 0·85 | Medium (feature importances available) |
| Naïve Bayes | 0·87 | 0·80 | 0·82 | Medium (independence assumption) |

All classifiers achieve AUC in the 0·87–0·93 range, confirming that the Lasso approach is not artificially advantaged and that the input feature space supports comparable discrimination across model families. Lasso was retained as the production model because it provides sparse, signed, directly interpretable coefficients that meet TRIPOD+AI interpretability expectations without sacrificing discrimination.

**Table S6. Colorado smoking status mapping rules**

Colorado smoking status was recorded as free text (14 unique entries). Deterministic keyword-based mapping to binary never/ever classification was performed as follows and audited by the senior author (CCH). Never-smoker (coded 0): “never” (n=182), “no” (n=2), “denies smoking” (n=1), “no smoking” (n=1), “no cigarette smoking” (n=1), “no history of smoking” (n=1), “non-smoker” (n=1). Ever-smoker (coded 1): “former” (n=267), “current” (n=38), “current some days” (n=5), “current every day” (n=2), “former pipe” (n=2), “former cigar” (n=1), “previous history of smoking” (n=1). A total of 505 records were mapped; no records had missing smoking data.

**Note on TRIPOD+AI checklist**

The complete TRIPOD+AI reporting checklist is provided as a separate Supplementary File (file 3 of the submission package).

**Figure S1. STARD participant flow**

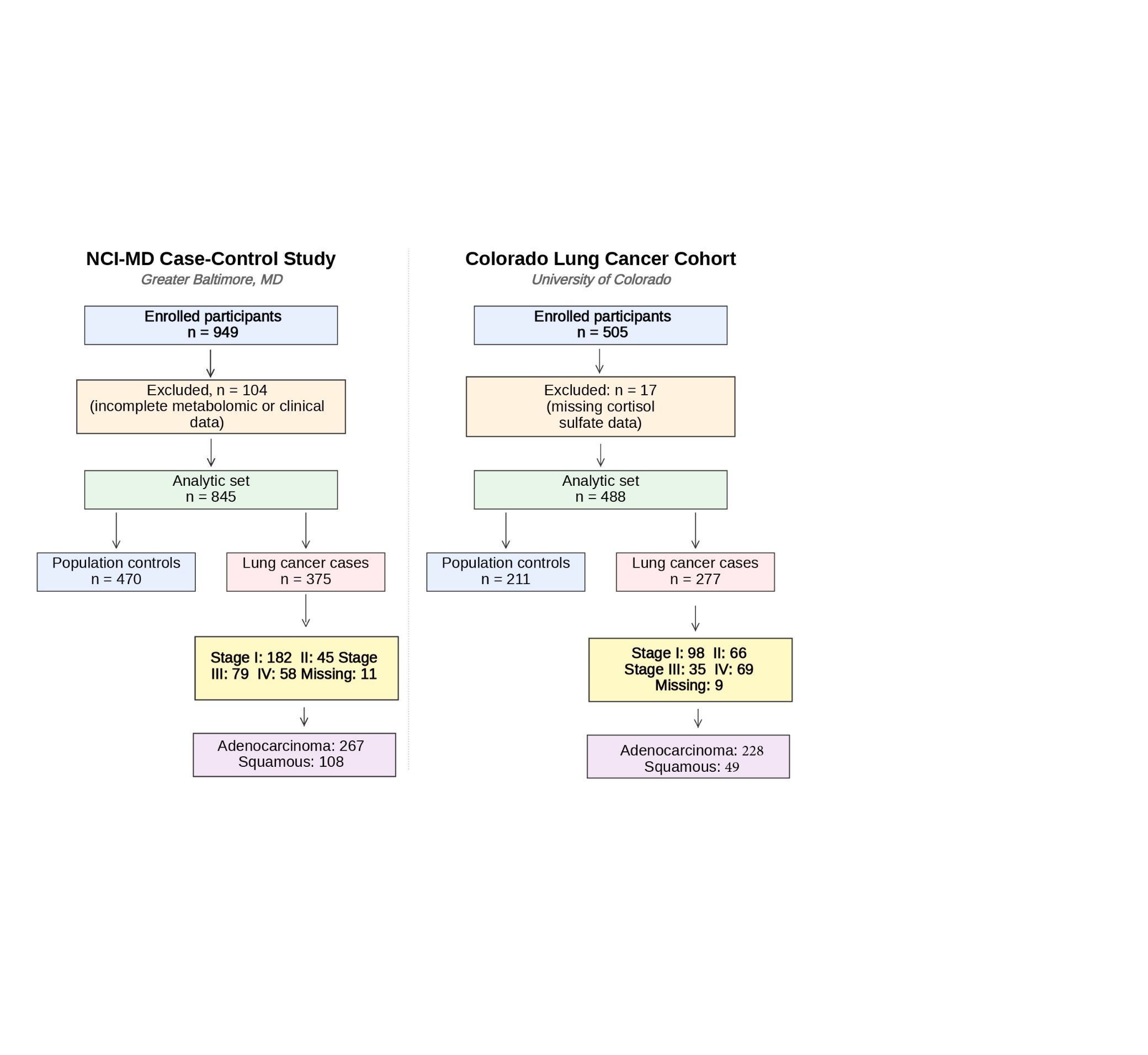

Figure S1. STARD-style participant flow diagrams for both cohorts. NCI-MD: 949 enrolled records → 845 complete-case analysis set after excluding 104 records with missing metabolomic or clinical data. Colorado: 505 enrolled records → 488 complete-case analysis set after excluding 17 records. Cases are stage I-IV; complete-case sample sizes are used throughout all analyses.

**Figure S2. uLCI predicted probability distribution across cancer stages**

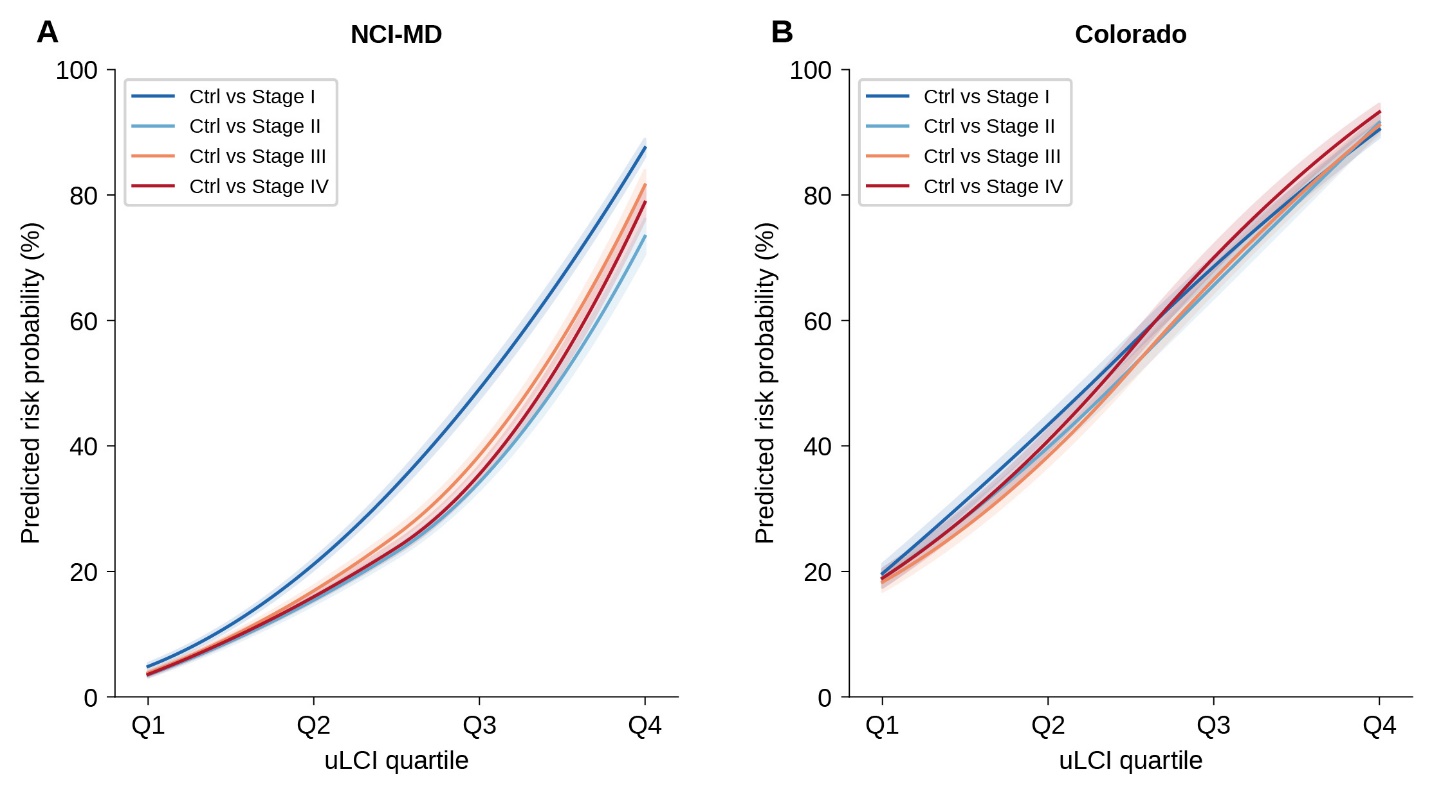

Figure S2. Distributions of uLCI predicted probabilities across cancer stages (I–IV) in both cohorts, shown as kernel density estimates restricted to cases only. (A) NCI-MD development cohort. (B) Colorado external validation cohort. The density view complements main Figure 1 by showing the full probability distribution shape within each stage category. uLCI distributions progressively shift toward higher values as stage advances, with stage III–IV cases concentrated above the Youden-J threshold (0·43, dotted line) and stage I cases showing the broadest distribution—consistent with the biological gradient of metabolic perturbation across disease progression.

**Figure S3. Overall, ROC curves**

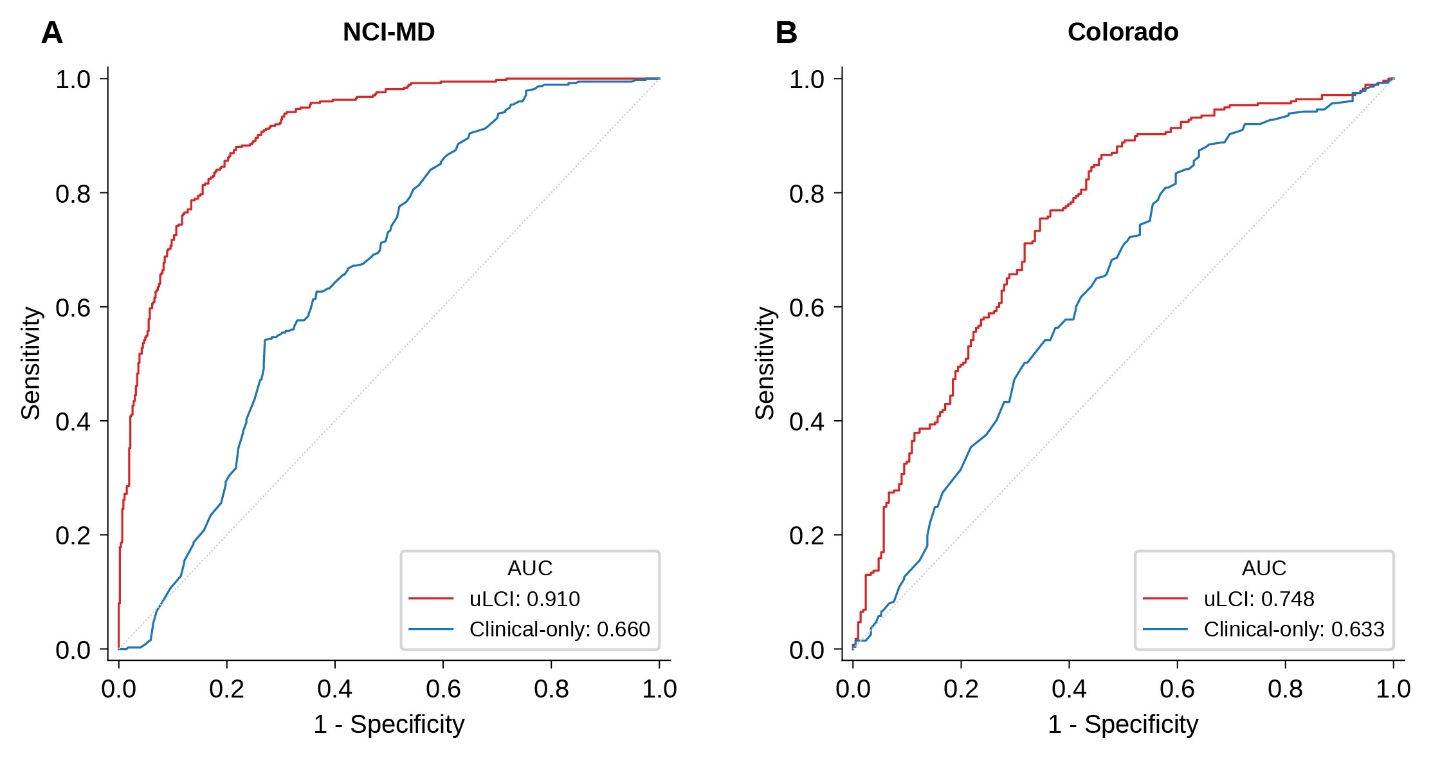

Figure S3. Overall receiver-operating-characteristic curves comparing uLCI (4 metabolites + 3 clinical variables) against a clinical-variables-only model. (A) NCI-MD development cohort, 10-fold cross-validation: uLCI AUC 0·906 (95% CI 0·887–0·926) versus clinical-only 0·632 (0·595–0·668). (B) Colorado independent external validation: uLCI AUC 0·748 (0·701–0·793) versus clinical-only 0·625 (0·581–0·669). The metabolite panel improves AUC by 0·27 in development and 0·12 in external validation (both bootstrap p<0·0001).

**Figure S4. uLCI by smoking status**

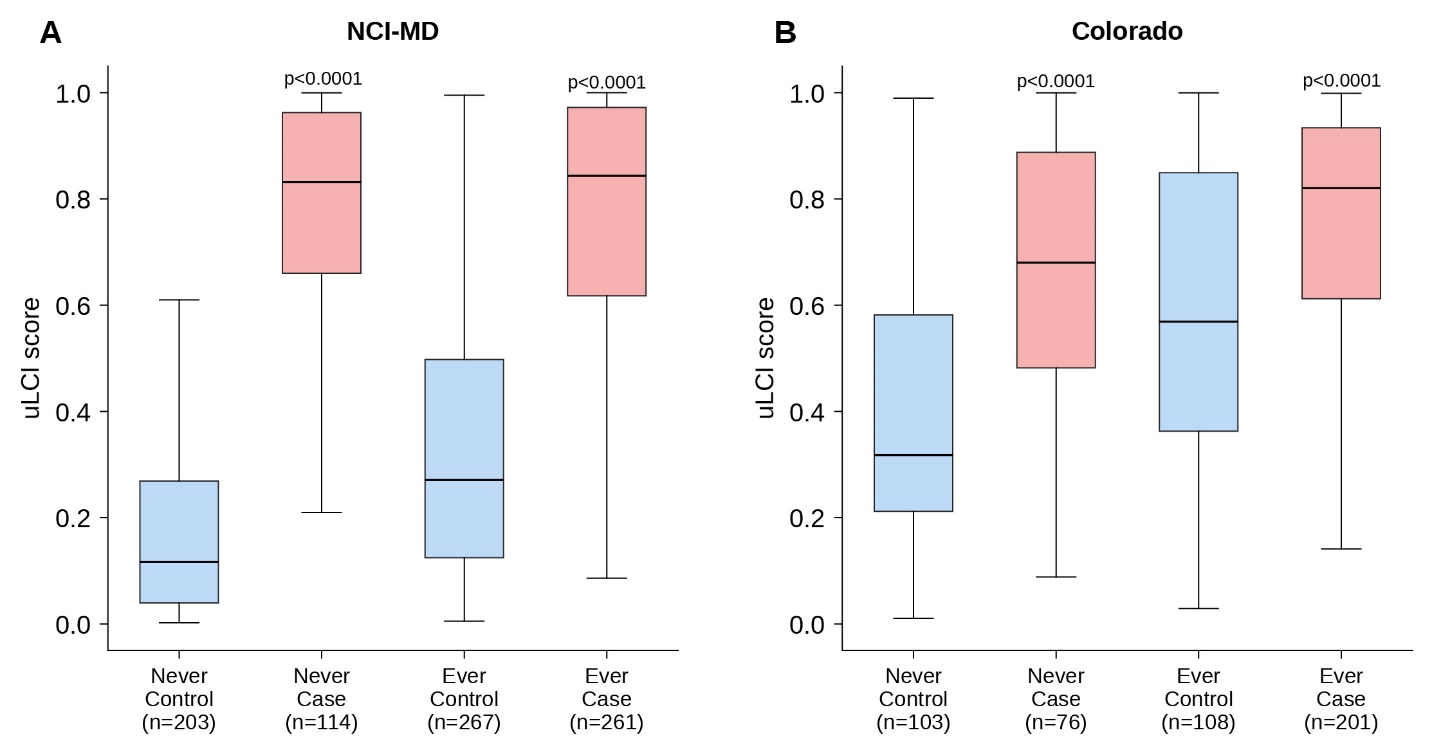

Figure S4. uLCI score distributions stratified by case-control status and smoking status. (A) NCI-MD development cohort. (B) Colorado external validation cohort. uLCI achieves significant case-control separation in both never-smokers and ever-smokers in both cohorts, indicating that the urinary metabolomic signal does not depend on smoking-induced metabolic perturbation and supporting potential applicability to populations currently outside LDCT-screening eligibility.

**Figure S5. Colorado calibration before and after recalibration**

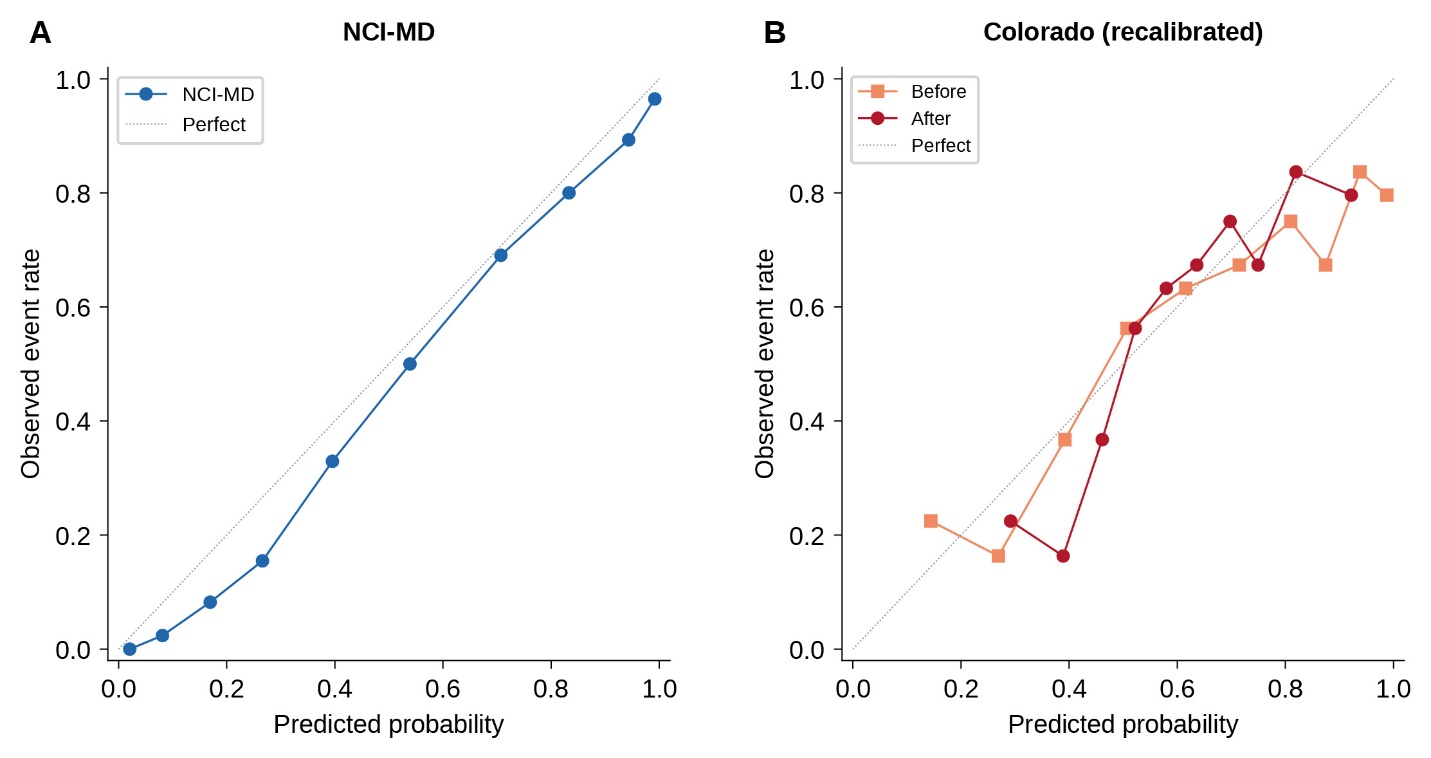

Figure S5. Colorado external validation calibration before and after intercept-slope recalibration. (A) Locked NCI-MD model applied to Colorado: Hosmer-Lemeshow p<0·001; Brier 0·206. (B) After Steyerberg-method recalibration (slope 0·524; intercept correction +0·075): Hosmer-Lemeshow p=0·033; Brier 0·203. AUC unchanged at 0·748 because intercept-slope recalibration is monotone and preserves rank-ordering.

**Figure S6. Overall survival by uLCI tertile — all stages combined**

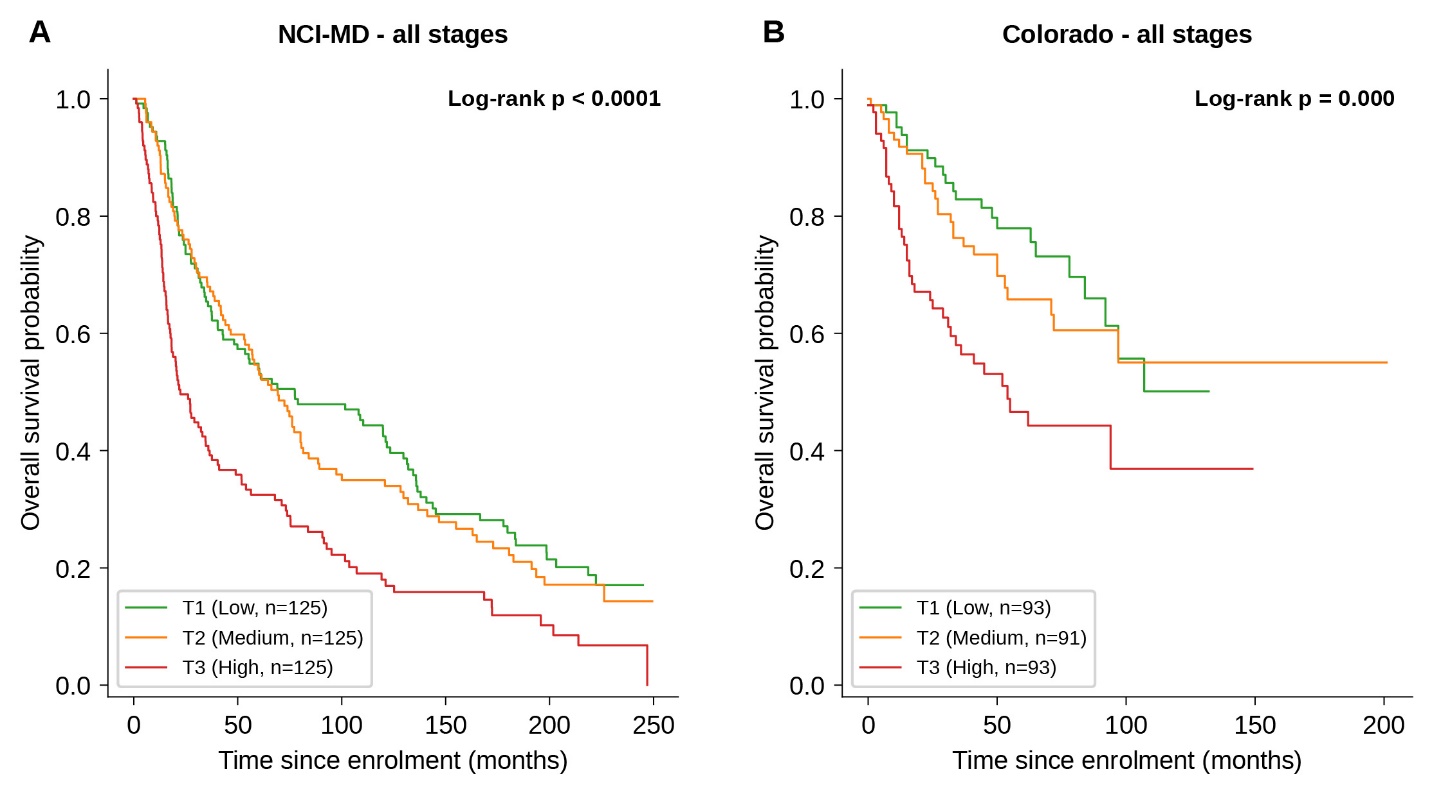

Figure S6. Kaplan–Meier overall-survival curves stratified by within-cohort uLCI tertile across all lung-cancer stages combined (I–IV). (A) NCI-MD cohort (n=375 with survival data; 297 events): 3-group log-rank p<0·0001. (B) Colorado cohort (n=277 with survival data; 91 events): 3-group log-rank p<0·0001. Tertile thresholds are within-cohort. uLCI stratifies survival across the full clinical stage spectrum, supporting its role as a quantitative marker of disease burden beyond binary case-control discrimination.

**Figure S7. Individual urinary metabolites — NCI-MD**

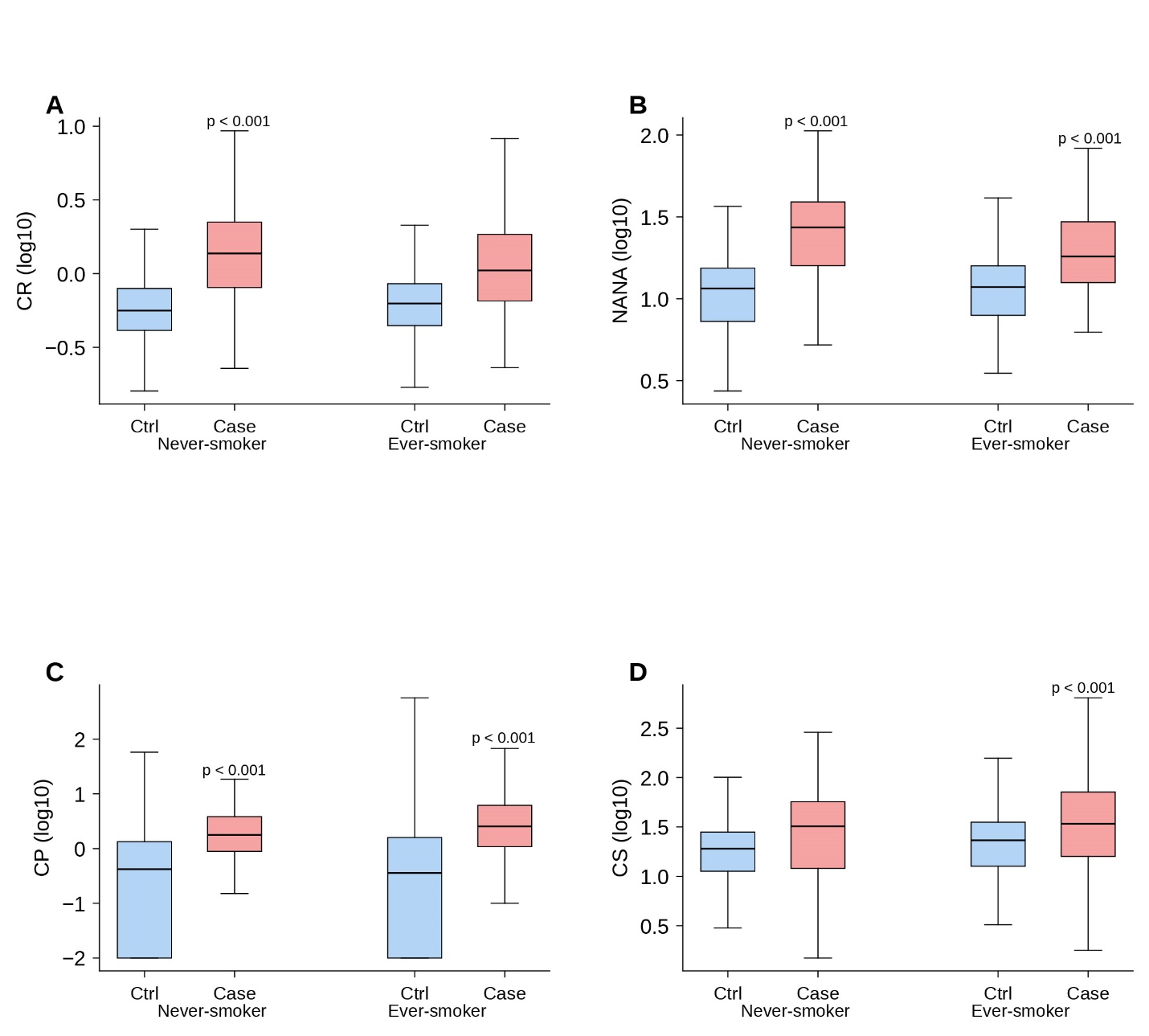

Figure S7. Distributions of the four urinary metabolites (CR, NANA, CP, CS) in the NCI-MD development cohort, stratified by smoking status. Boxplots show log10-transformed, creatinine-normalised levels in controls versus cases. Wilcoxon rank-sum p-values shown above each stratum (red indicates p<0·05). All four metabolites are significantly elevated in cancer cases relative to controls in both smoking strata. Two of these metabolites (CR and NANA) extend our previously published two-metabolite work; CP and CS are extensions of the panel.

**Figure S8. Individual urinary metabolites — Colorado**

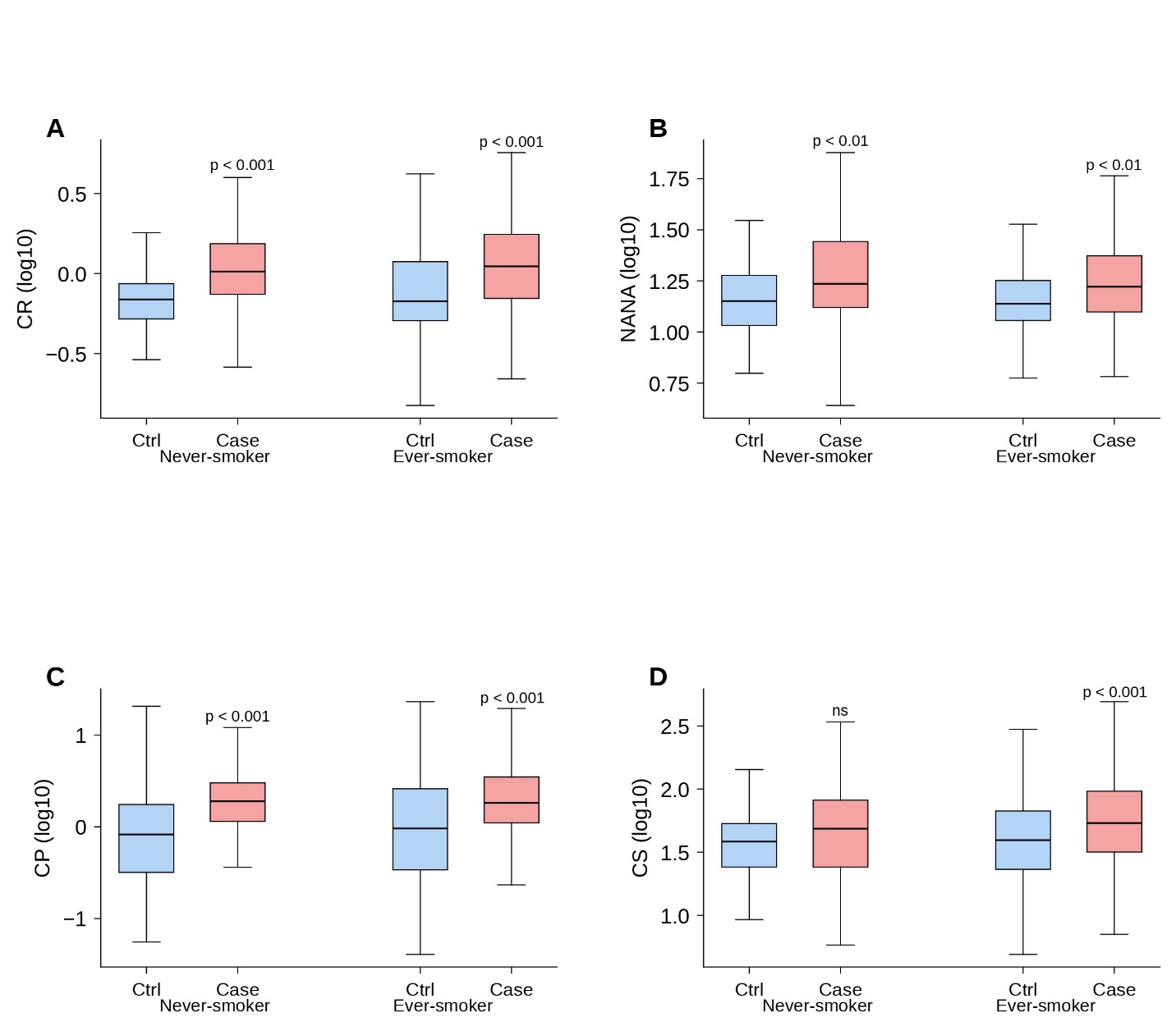

Figure S8. Distributions of the four urinary metabolites in the Colorado external validation cohort, stratified by smoking status. Boxplots show log10-transformed, creatinine-normalised levels in controls (n=211) versus cases (n=277, stages I-IV). All four metabolites replicate the direction of effect observed in NCI-MD with attenuated effect sizes, consistent with the broader stage distribution and population shift in Colorado.
