## Supplementary material for "The urinary-metabolite-based lung cancer index (uLCI): an interpretable machine-learning risk model for early-stage disease": Completed TRIPOD+AI reporting checklist

*Supplementary File 1 — Reporting compliance for:*

**"The urinary-metabolite-based lung cancer index (uLCI): an interpretable machine-learning risk model for early-stage disease"**

*Khan MA, Pine SR, Gonzalez FJ, Wang XW, Harris CC, Patel DP*

This checklist follows the TRIPOD+AI statement (Collins GS, Moons KGM, Dhiman P, et al. BMJ 2024;385:e078378). Items are scored as "D" (development), "E" (evaluation/external validation), or "D;E" (both). uLCI is a prediction model that was both developed (NCI-MD cohort) and externally evaluated (Colorado cohort); the manuscript reports both D and E aspects.

For each item, "Location" indicates the section of the main manuscript where the item is addressed: T = Title; A = Abstract; I = Introduction; M = Methods; R = Results; D = Discussion; S = Supplement.

| **Item** | **TRIPOD+AI checklist item** | **Response — uLCI manuscript** | **Location** |
| --- | --- | --- | --- |
| **TITLE** | | | |
| **1** | Identify the study as developing or evaluating the performance of a multivariable prediction model, the target population, and the outcome to be predicted. | Title states "The urinary-metabolite-based lung cancer index (uLCI): an interpretable machine-learning risk model for early-stage disease." Identifies the model (urinary metabolite risk index), target population (early-stage lung cancer), and outcome (case-control discrimination). Brand name (uLCI) introduced in abstract. | T |
| **ABSTRACT** | | | |
| **2** | Structured abstract addressing each item of the TRIPOD+AI for Abstracts checklist. | Structured abstract reports background, methods (data sources, predictors, sample size, statistical methods), findings (discrimination, calibration, decision-curve, subgroups), and interpretation. | A |
| **INTRODUCTION** | | | |
| **3a** | Explain healthcare context (diagnostic or prognostic) and rationale for developing or evaluating the prediction model, including references to existing models. | Diagnostic context (early-stage lung adenocarcinoma detection). LDCT, PLCOm2012, DELFI, Galleri described as existing approaches with stated limitations. Prior 2-metabolite work (Dalal 2024, Haznadar 2016) referenced as direct foundation. | I |
| **3b** | Describe target population and intended purpose of the prediction model in the care pathway, including its intended users. | Target population: individuals at risk of early-stage lung adenocarcinoma, including never-smokers excluded from LDCT eligibility. Intended purpose: non-invasive triage tool to elevate pre-test probability for confirmatory imaging. Intended users: clinicians and screening programmes. | I |
| **3c** | Describe any known health inequalities between sociodemographic groups. | Disparities in lung cancer outcomes by race (African American populations) and by smoking status (never-smokers excluded from LDCT) are explicitly described, with cited references. | I |
| **4** | Specify the study objectives, including whether the study describes development or validation (or both). | Stated explicitly: development in NCI-MD cohort and independent external validation in Colorado cohort under a locked, non-refitted model. | I, end |
| **METHODS — Data and participants** | | | |
| **5a** | Describe sources of data separately for development and evaluation datasets, rationale, and representativeness. | NCI-MD case-control study (development) and Colorado Lung Cancer Cohort (evaluation), both described separately with institutional setting, recruitment, and representativeness. Both are prospectively-collected biobanked cohorts. | M |
| **5b** | Specify dates of collected participant data, including start and end of accrual; if applicable, end of follow-up. | NCI-MD enrollment from early 2000s; Colorado prospectively biobanked. Specific accrual dates available in cohort source publications and stated in Methods. | M |
| **6a** | Specify key elements of the study setting (e.g., primary care, secondary care), including the number and location of centres. | NCI-MD: greater Baltimore metropolitan region, single primary recruitment centre (NCI). Colorado: University of Colorado, single primary recruitment centre. Both tertiary academic centres. | M |
| **6b** | Describe eligibility criteria for study participants. | NCI-MD: histologically confirmed lung adenocarcinoma cases plus population-based frequency-matched controls. Colorado: histologically confirmed lung cancer cases plus controls. Specific exclusion: missing metabolomic or clinical variables. | M |
| **6c** | Give details of any treatments received, and how handled, if relevant. | Urine samples obtained prior to surgical resection or systemic therapy initiation. Pre-treatment status confirmed in Methods. | M |
| **7** | Describe any data pre-processing and quality checking, including whether similar across sociodemographic groups. | Metabolite values creatinine-normalised. Free-text smoking categories in Colorado mapped to binary never/ever schema by deterministic keyword rule with audit by senior authors. Pre-processing applied uniformly across racial subgroups; no group-specific cleaning. | M |
| **METHODS — Outcome and predictors** | | | |
| **8a** | Clearly define the outcome that is being predicted, time horizon, and the rationale, and whether method of outcome assessment is consistent across sociodemographic groups. | Primary outcome: lung adenocarcinoma case status (binary, histologically confirmed). Time horizon: not time-to-event for primary discrimination; survival outcomes assessed separately (overall survival post-resection). Histological confirmation consistent across racial and smoking subgroups. | M |
| **8b** | If outcome assessment requires subjective interpretation, describe the qualifications and demographic characteristics of outcome assessors. | Histological diagnosis by board-certified pathologists at each institution per AJCC staging guidelines. Not subject to inter-rater variability concerns at this level. | M |
| **8c** | Report any actions to blind assessment of the outcome. | Outcome (case-control status) determined at clinical enrollment, prior to metabolomic measurement. Metabolomic measurements were performed blinded to case-control status (lab batch processing). | M |
| **9a** | Describe the choice of initial predictors and any pre-selection before model building. | Four urinary metabolites (CR, NANA, CP, CS) prespecified based on prior published work (Mathé 2014, Haznadar 2016, Dalal 2024). Three clinical variables (age, race, smoking status) prespecified from epidemiological evidence. Additional candidates (sex, pack-years, BMI, alcohol, occupation, diabetes) screened and not retained. | M |
| **9b** | Clearly define all predictors, including how and when measured (and any blinding). | CR, NANA, CP, CS quantified by targeted LC-MS/MS with stable-isotope internal standards. MRM transitions, retention times, and IS concentrations detailed in Supplementary Table S2. Predictors measured before treatment initiation, blinded to case-control status. | M, S |
| **9c** | If predictor measurement requires subjective interpretation, describe qualifications/demographics of assessors. | Not applicable — LC-MS/MS quantification is fully automated with internal-standard normalisation; no subjective interpretation. | M |
| **METHODS — Sample size and missing data** | | | |
| **10** | Explain how the study size was arrived at, justify sufficiency to answer the research question. Include sample size calculation details. | Sample size determined by availability of complete metabolomic and clinical data in both biobanked cohorts (NCI-MD n=845; Colorado n=488). Both cohorts comfortably exceed the events-per-variable rule (≥20 outcomes per candidate predictor); 7 candidate predictors required ≥140 outcomes minimum, observed 375 in NCI-MD and 277 in Colorado. | M |
| **11** | Describe how missing data were handled. Provide reasons for omitting any data. | Complete-case analysis. Of 949 NCI-MD records, 845 had complete data for all four metabolites and three clinical variables (104 omitted for missingness). Of 505 Colorado records, 488 had complete data (21 omitted, predominantly for indeterminate disease status or missing metabolomics). Reasons reported in Results flow text. | M, R |
| **METHODS — Analytical methods** | | | |
| **12a** | Describe how the data were used (for development and evaluation), including partitioning and sample size considerations. | NCI-MD: 10-fold cross-validation for unbiased internal performance estimation, with cross_val_predict ensuring no observation contributed to fitting its own predicted probability. Colorado: locked-model external evaluation with no re-fitting, no hyperparameter re-tuning, and no threshold re-selection. No data leakage between development and evaluation. | M |
| **12b** | Describe how predictors were handled in the analyses (functional form, rescaling, transformation). | Metabolite values log-transformed (log1p) and creatinine-normalised. All features standardised to zero mean and unit variance using NCI-MD parameters and applied identically to Colorado. Age continuous, race binary (AA vs EA), smoking binary (never vs ever). | M |
| **12c** | Specify the type of model, rationale, all model-building steps including any hyperparameter tuning, and method for internal validation. | L1-regularised (Lasso) logistic regression. Rationale: produces sparse, interpretable coefficient vectors well-suited to compact biomarker panels and modest-dimensional tabular data; provides implicit feature selection while limiting overfitting. Hyperparameter (regularisation strength λ) selected across 20 candidate values by 10-fold cross-validated AUC. Benchmark comparison with k-NN, SVM (RBF), random forest, naïve Bayes under identical CV (Supplementary Table S4). | M, S |
| **12d** | Describe handling of heterogeneity in parameter estimates across clusters. | Two independent cohorts treated as separate sources rather than pooled, in line with TRIPOD-Cluster recommendations. Subgroup analyses by race, sex, and smoking status reported with bootstrap CIs. | M, R |
| **12e** | Specify all measures and plots used to evaluate model performance (discrimination, calibration, clinical utility) and to compare multiple models. | Discrimination: AUC with 2,000-iteration bootstrap 95% CI, sensitivity, specificity, PPV, NPV, Cohen's κ, accuracy, false-positive rate at the Youden-J threshold. Calibration: Hosmer-Lemeshow test (10 deciles), calibration plot (10 quantile bins), Brier score. Clinical utility: decision-curve analysis. Pairwise model comparison: bootstrap-derived AUC differences. | M, R |
| **12f** | Describe any model updating (e.g., recalibration) arising from model evaluation. | Intercept-slope recalibration performed on Colorado data (Steyerberg method). Calibration slope 0·524 and intercept correction +0·075. Hosmer-Lemeshow improved from p<0·001 (uncalibrated) to p=0·033 (recalibrated). Rank-ordering and AUC unchanged. Recalibration framed as a routine pre-deployment step. | M, R |
| **12g** | For model evaluation, describe how predictions were calculated (formula, code, object, API). | Locked Lasso logistic regression model: uLCI = σ(β·X + β₀), with σ the logistic sigmoid. Coefficients reported in Supplementary Table S3. Analytical code prepared for deposit on Zenodo. | M, S |
| **13** | If class imbalance methods were used, state why and how, and any subsequent recalibration. | Not applicable — case-control prevalence approximately balanced in both cohorts (NCI-MD 44% cases; Colorado 57% cases). No imbalance-correction methods (e.g., SMOTE, under/over-sampling) applied. | M |
| **14** | Describe approaches to address model fairness and their rationale. | Race included as a model input (binary AA vs EA) to allow the model to learn population-specific patterns. Subgroup performance reported separately for African American and European American participants. NCI-MD development cohort intentionally diverse (34% African American). Colorado validation cohort had only n=18 African American participants, limiting fairness evaluation in validation; flagged as a key limitation requiring replication in larger diverse cohorts. | M, R, D |
| **15** | Specify the output of the prediction model (probabilities, classification). Provide rationale for any classification and how thresholds were identified. | Primary output: predicted probability ∈[0,1]. Binary classification at Youden-J threshold optimised on NCI-MD cross-validated predictions (threshold = 0·43) and applied identically to Colorado without re-selection. Tertile thresholds reported for prognostic Kaplan-Meier analysis. | M, R |
| **16** | Identify any differences between development and evaluation data in setting, eligibility, outcome, predictors. | Differences acknowledged explicitly: stage distribution (NCI-MD stage I-II only by design; Colorado stages I-IV); racial composition (NCI-MD 34% AA; Colorado 4% AA); free-text smoking variable in Colorado (mapped to binary). These are reported in Methods and discussed as contributors to the development-vs-validation AUC gap. | M, D |
| **17** | Name the institutional research board or ethics committee that approved the study and describe informed consent. | NCI-MD: NIH/NCI Institutional Review Board approval; written informed consent obtained from all participants. Colorado: University of Colorado Institutional Review Board approval; written informed consent obtained. | M |
| **OPEN SCIENCE** | | | |
| **18a** | Give the source of funding and the role of the funders for the present study. | Intramural Research Program, Center for Cancer Research, National Cancer Institute, NIH. Funder had no role in study design, data collection, analysis, interpretation, or writing. | Funding |
| **18b** | Declare any conflicts of interest and financial disclosures for all authors. | Authors declare no competing financial interests directly relevant to this work. NCI Technology Transfer Office is exploring licensing pathways for the analytical assay; not active during data collection. | Declarations |
| **18c** | Indicate where the study protocol can be accessed or state that a protocol was not prepared. | No formal pre-registered protocol for the analytical extension was deposited; analysis plan documented internally and presented to senior authors prior to model fitting. Original NCI-MD and Colorado cohort protocols approved by respective IRBs. | M |
| **18d** | Provide registration information for the study or state that it was not registered. | Study not registered in a public registry (typical for biomarker development studies using retrospective biobanked samples). Cohort source studies are NIH-sponsored and publicly described. | M |
| **18e** | Provide details of the availability of the study data. | De-identified individual participant data available to bona fide researchers on reasonable request to the corresponding authors, subject to NCI IRB approval and a data-use agreement. Data dictionary prepared for distribution. | Data sharing |
| **18f** | Provide details of the availability of the analytical code. | R 4·2·0 analytical code (data harmonisation, Lasso fitting, cross-validation, external validation, bootstrap CI estimation, calibration analysis, recalibration, decision-curve analysis, permutation importance, survival analysis, figure generation) deposited on Zenodo upon acceptance with DOI assigned. Software versions: R 4·2·0, glmnet 4·1, pROC 1·18, caret 6·0, ResourceSelection 0·3, dcurves 0·4, nricens 1·6, survival 3·5, survminer 0·4, ggplot2 3·5. | Data sharing |
| **PATIENT AND PUBLIC INVOLVEMENT** | | | |
| **19** | Provide details of any patient and public involvement during the design, conduct, reporting, interpretation, or dissemination of the study. | No patient or public partners were directly involved in the design or conduct of this retrospective biomarker analysis. Patient advocacy organisations (LUNGevity, GO2 Foundation) consulted historically on clinical priorities for never-smoker lung cancer detection at the parent-cohort level. Prospective evaluation will involve formal PPI consultation. | Acknowledgements |
| **RESULTS** | | | |
| **20a** | Describe the flow of participants through the study, including the number with and without the outcome and follow-up time. A diagram may help. | Participant flow described in Methods text. NCI-MD: 949 records → 845 with complete data (470 controls, 375 cases). Colorado: 505 records → 488 with complete data (211 controls, 277 cases). STARD-style flow diagram in Supplementary Figure S1. | M, R, S |
| **20b** | Report the characteristics overall and for each data source: key dates, key predictors, treatments, sample size, outcome events, follow-up, missing data. | Table 1 reports cohort baseline characteristics: total n, controls/cases, stage distribution, smoking status, age (mean, SD), sex, race. Subgroup counts reported. | Table 1 |
| **20c** | For model evaluation, show a comparison with development data of the distribution of important predictors and outcome. | Distribution of all four metabolites and clinical variables reported separately for NCI-MD (Figure 1) and Colorado (Figure 2). Differences in age, race composition, and stage distribution between cohorts described in Methods and Discussion. | Figs 1, 2; Table 1 |
| **21** | Specify the number of participants and outcome events in each analysis. | Each analysis (overall, never-smoker, ever-smoker, AA, EA, stage I, stage II) reports its n and event count, including in subgroup table (Table 1) and figure captions. | R, Table 1 |
| **RESULTS — Model development and evaluation** | | | |
| **22** | Provide the full prediction model (i.e., all the regression coefficients, or alternative output, including baseline survival) to allow predictions on new individuals and for independent evaluation of the model. | Full uLCI model: Lasso coefficients on standardised features (CR β=+1·27, NANA β=+1·25, CP β=+0·98, CS β≈0, age β=-0·24, race-AA β=-0·55, smoking β=+0·39, intercept β₀); standardisation parameters reported in Supplementary Table S3; sigmoid linkage explicit in Methods. | M, S, Table S3 |
| **23** | Report model performance estimates with 95% CIs, separately for development and evaluation. If multiple models, report performance for each. | Bootstrap 95% CIs (2,000 iterations) reported for every discrimination metric. NCI-MD: AUC 0·906 (CI 0·887-0·926); Colorado: AUC 0·748 (CI 0·701-0·793). Benchmark classifiers reported in Supplementary Table S4. | R, Table 1 |
| **24** | If risk groups created, provide group details and number of participants and outcome events in each. | Tertile risk grouping reported for survival analysis: low/medium/high uLCI tertiles in each cohort. Tertile boundaries derived from within-cohort quantiles; sample sizes per tertile reported in Figure 3 and text. | R, Fig 8 |
| **25** | Report results from any subgroup or sensitivity analysis. | Subgroup performance reported in Table 1 (smoking, race, stage). Sensitivity analyses include benchmark classifiers (Supplementary Table S4), recalibration (Methods, Results), and nested-model comparison (Table 1). | R, Table 1, S |
| **DISCUSSION** | | | |
| **26** | Give an overall interpretation of the main results, including issues of fairness in the context of the objectives and previous studies. | Discussion opens with stage I-II performance interpretation. Fairness explicitly addressed: cross-racial preservation of AUC in NCI-MD (AA 0·91, EA 0·89); limitation explicitly noted regarding small AA n in Colorado. | D |
| **27** | Discuss any limitations of the study (e.g., non-representative sample, sample size, overfitting, missing data) and effects on biases, statistical uncertainty, and generalisability. | Six limitations explicitly discussed: (i) case-control rather than prospective screening design; (ii) absence of head-to-head comparison with DELFI/Galleri on same samples; (iii) Colorado calibration miscalibration with recalibration shown; (iv) racial composition mismatch between cohorts; (v) free-text smoking categorisation noise; (vi) CP biological mechanism the subject of a separate manuscript. | D |

**TRIPOD+AI for Abstracts**

TRIPOD+AI for Abstracts is a separate, shorter checklist for the structured abstract. Reporting items are tabulated below.

| **Item** | **TRIPOD+AI checklist item** | **Response — uLCI manuscript** | **Location** |
| --- | --- | --- | --- |
| **A1** | Identify the study as developing or evaluating the performance of a multivariable prediction model, the target population, and the outcome to be predicted. | Abstract Background: "We aimed to develop and externally validate the urinary metabolite Lung Cancer Index (uLCI) ... for non-invasive detection of stage I-II lung adenocarcinoma." | A — Background |
| **A2** | Briefly describe the data sources used. | NCI-MD case-control cohort (n=845) and Colorado Lung Cancer Cohort (n=488) named explicitly. | A — Methods |
| **A3** | State the predictors and outcome. | Four urinary metabolites (CR, NANA, CP, CS) + three clinical variables (age, race, smoking status) named. Outcome: lung adenocarcinoma case-control status. | A — Methods |
| **A4** | Specify the model type and statistical methods. | Lasso-regularised logistic regression with 10-fold cross-validation. Bootstrap 95% CIs. Calibration analysis. Decision-curve analysis. | A — Methods |
| **A5** | Specify the sample size and number of outcome events. | NCI-MD: 470 controls, 375 cases. Colorado: 211 controls, 277 cases. Both reported in Methods. | A — Methods |
| **A6** | State the main results (discrimination and calibration) with confidence intervals. | AUC 0·906 (95% CI 0·887-0·926) NCI-MD, 0·748 (0·701-0·793) Colorado. Subgroup AUCs reported. Calibration before/after recalibration reported. | A — Findings |
| **A7** | State an interpretation of the findings. | Interpretation paragraph positions uLCI as non-invasive triage tool with externally validated performance for stage I-II adenocarcinoma. | A — Interpretation |
| **A8** | Funding source and registration (if relevant). | NIH Intramural Research Program. Not registered (retrospective biomarker analysis using biobanked samples). | A — Funding |

Official checklist and supplementary materials available at: www.tripod-statement.org

This completed TRIPOD+AI checklist accompanies the main manuscript "The urinary-metabolite-based lung cancer index (uLCI): an interpretable machine-learning risk model for early-stage disease" (Khan, Mathé, Wang, Harris, Patel). All 27 numbered items of TRIPOD+AI are addressed in the manuscript text, supplementary materials, or analytical code repository. Items rated as "not applicable" (e.g., subjective interpretation, class imbalance methods) are justified.
